# Death, Demography and the Denominator: Age-Adjusted Influenza-18 Mortality in Ireland^*^

**DOI:** 10.1101/2020.05.26.20113381

**Authors:** Christopher L. Colvin, Eoin McLaughlin

## Abstract

Using the Irish experience of the 1918-1919 Spanish flu pandemic (“Influenza-18”), we demonstrate that pandemic mortality statistics are sensitive to the demographic composition of a country. We build a new spatially disaggregated population database for Ireland’s 32 counties for 1911-1920 with vital statistics on births, ageing, migration and deaths. Our principal contribution is to show why, and how, age-at-death data should then be used to construct the age-standardised statistics necessary to make meaningful comparisons of mortality across time and space. We conclude that studies of the economic consequences of pandemic pandemics must better control for demographic factors if they are to yield useful policy-relevant insights. For example, while Northern Ireland had a higher crude death rate during the first wave of the Covid-19 pandemic, it also has an older population; age-adjusted mortality paints a very different picture.

## 1. Introduction

Lessons from the Great Depression were widely employed by economists and policymakers to understand and respond to the Great Recession (Eichengreen 2012). As the world’s last truly global pandemic before Covid-19, the influenza pandemic of 1918-1919 (“Influenza-18”) is now being used in much the same way. A plethora of new studies has already emerged which use this historical pandemic to discuss the potential short- and medium-term social, economic and political ramifications of Covid-19 (see, e.g., Barro et al. 2020; Basco et al. 2020; Benmelech and Boberg-Fazlic et al. 2020; Frydman 2020; Carillo and Jappelli 2020; Chapelle 2020; Correia et al. 2020; Dahl et al. 2020; Lilley et al. 2020; Lin and Meissner 2020; Velde 2020).

But learning lessons from history is hard, and the history profession has long shied away from doing so out of fear that practically anything can be justified by appealing to one or other interpretation of past events (Colvin and Winfree 2019). The advantage economists had in 2008 was that we knew a lot about the Great Depression; indeed, the field of macroeconomics ostensibly originated in that very crisis. We do not have the same home advantage when we look to Influenza-18 to draw our lessons; economists are relatively new to this topic and run the risk of making rookie errors which distort our findings and could lead to poorly designed policy advice.

We highlight one such error in a case study of Ireland, which in 1918 was still an impoverished region of one of the world’s most advanced economies: the United Kingdom of Great Britain and Ireland. We show that fragile population estimates combined with a failure to adjust mortality statistics for age-at-death have distorted our understanding of this historical pandemic. Just as currencies need to be adjusted for purchasing power when comparing the output of different economies, mortality statistics need to undergo age standardisation when comparing the impact of pandemics across different populations.

Recent debates in the *Proceedings of the National Academy of Sciences* and *Science Magazine* call for greater use of demographic methods in the study of Covid-19’s mortality burden (Dowd et al. 2020a; Nepomuceno et al. 2020; Dowd et al. 2020b; Nordling 2020). Meanwhile, medical scholars are reminding their peers about the importance of measuring excess mortality rather than cause-specific mortality when gauging the impact of Covid-19 (e.g., Beaney et al. 2020). The economics community now needs to take stock of these and related discussions. Some models of the macroeconomic impact of pandemics already incorporate historical epidemiology (e.g., Keogh-Brown et al. 2010a,b). They should now also make adjustment for demographic change. In this paper, we prescribe off-the-shelf solutions from the field of demography, which can be easily employed in economic research into Influenza-18 and Covid-19 alike.

## 2. Why Demography Matters

Influenza-18 was a highly infectious virus with significant human and economic costs. The Spanish flu, as it is still commonly known, may have had a global death toll upward of 50 million (Jester et al. 2019; Jordan et al. 2019).^1^ An estimated 2.6 million deaths occurred in Europe alone – 1.1 per cent of the continent’s population (Ansart et al. 2009). While their ultimate cause of death is generally thought to have been the influenza A virus subtype H1N1, typical proximate causes of death during this pandemic were secondary bacterial pneumonia infections and respiratory failure (Taubenberger 2006) – much as in the case of SARS-CoV-2 (ECDC 2020).

<u>Figure 1</u> plots the crude mortality rate of the constituent polities of the UK, alongside that of other countries, at an annual frequency between 1900 and 1920.^2^ <u>Figure 2</u> reports crude excess mortality for Ireland at a quarterly frequency across the same period. These two figures reveal the severity of the pandemic, but they also highlight the fact that Influenza-18 took place within a high mortality environment where outbreaks and epidemics were more commonplace than today. Indeed, this was a time of epidemiological transition, when infectious (exogenous) diseases remained a more significant cause of death than chronic lifestyle (endogenous) causes (Omran 1971; Dyson 2010).

**Figure 1:**
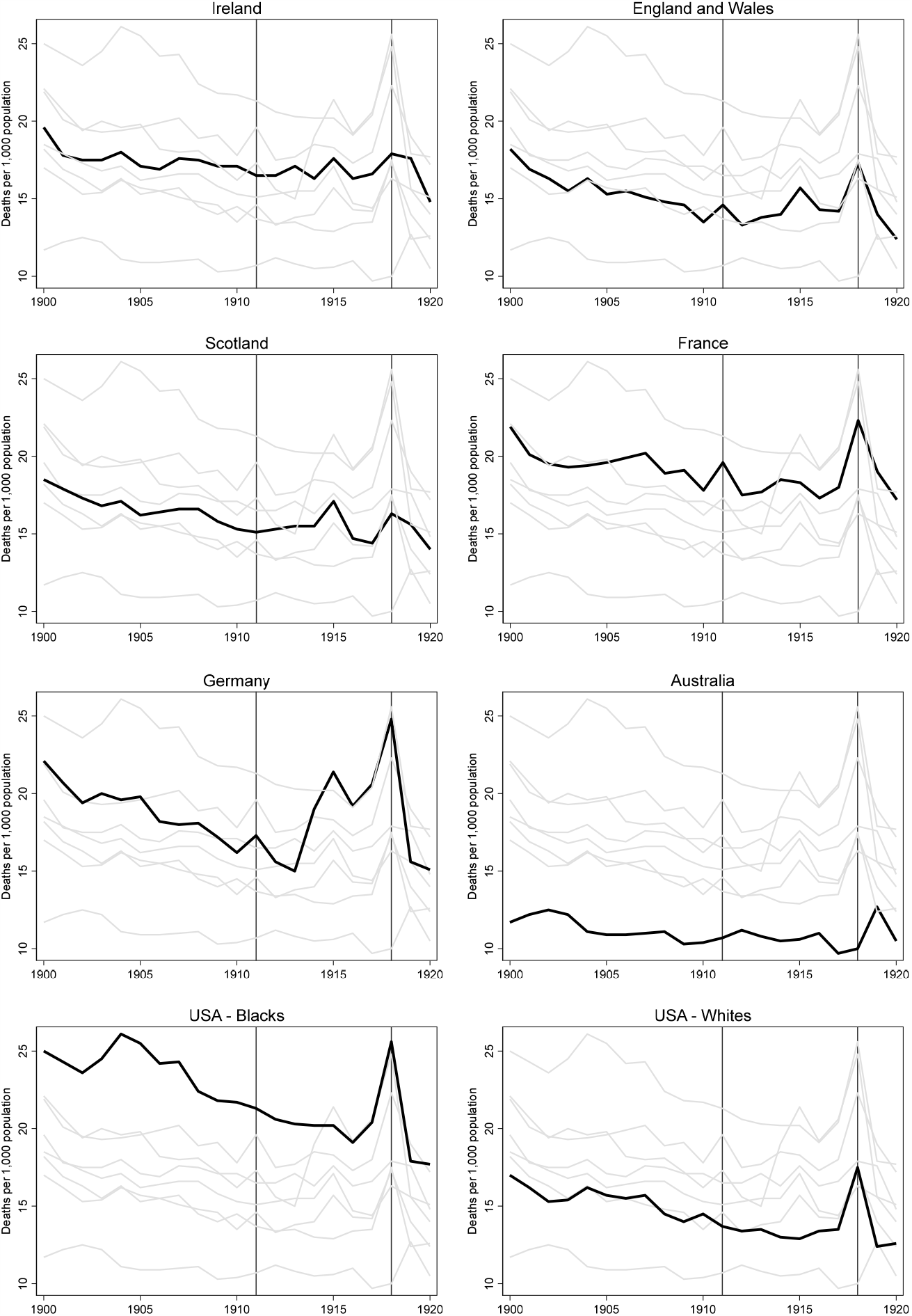
Crude mortality rates in Ireland and other countries (1900-1920, annual frequency) Note: 1911 and 1918 are indicated with vertical lines. US data are reported by ethnicity only. Source: Mitchell (2013).

**Figure 2:**
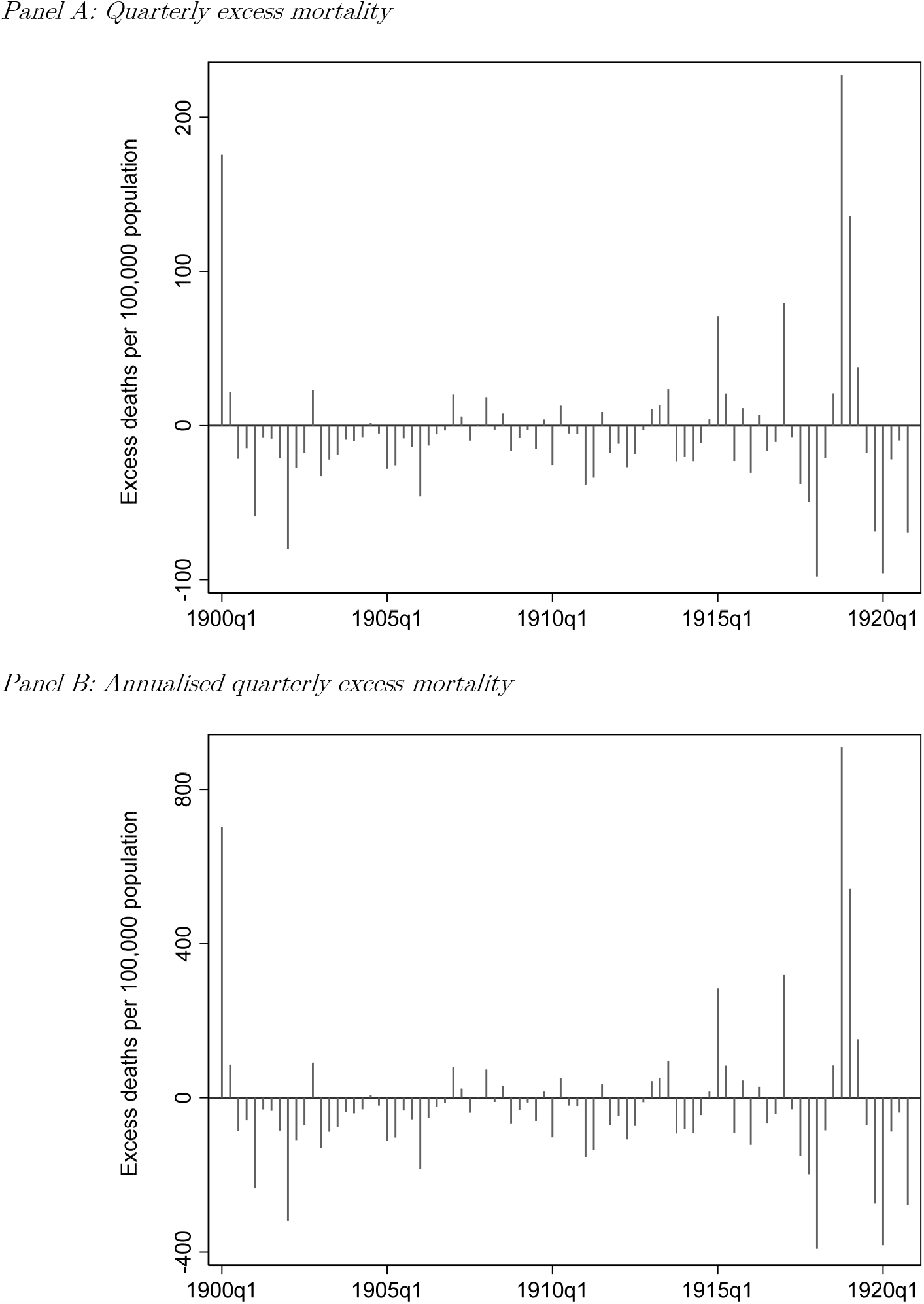
Crude excess mortality rates in Ireland (1900-1920) Note: Quarterly excess mortality is defined here as current mortality (*t*_*qx*_) minus the average mortality of the same quarter in the previous four years (*t*− 1_*qx*_, *t*− 2_*qx*_, *t*− 3_*qx*_, *t*− 4_*qx*_). For example, excess mortality for 1900q1 compares 1900q1 with the average of 1899q1, 1898q1, 1897q1 and 1896q1. Annualised quarterly excess mortality permits us to make comparisons with annual mortality rates, and are quarterly crude mortality multiplied by four. Source: BPP (1907) and BPP (1915b).

Influenza-18 originated from an unknown source (Taubenberger et al. 2001), and typically arrived in countries in major trading ports – carried, among others, by military personnel. The disease diffused through populations in a process of spatial contagion along major transportation infrastructure, typically in multiple waves (Smallman-Raynor et al. 2002). Great Britain and Ireland saw three waves of disease, with the second being the deadliest (Honingsbaum 2009; Milne 2018). But while all areas of the world saw excess mortality, rates differed significantly across countries (Murray et al. 2006); Influenza-18 proved particularly deadly in developing countries (Chandra et al. 2012). There was also significant heterogeneity in the flu’s health impact within countries, an outcome driven by local differences in demography, density, economy, environment and policy (Patterson and Pyle 1991; Hatchett et al. 2007; Clay et al. 2018, 2019).

Age mattered. Indeed, worldwide the conventional wisdom is Spanish flu was particularly fatal to those aged between 20 and 40 years (Johnson and Mueller 2002). One hypothesis is these young adults were immunologically naïve, with older age groups benefitting from inbuilt immunity due to expose to previous influenza outbreaks (Palese 2004; Taubenberger 2006). We can also speculate that young adults, who are more likely to be in employment, are inherently more at risk of catching the disease in societies which do not implement “lockdown” measures (cf. Hatchett 2007). Sex also mattered; men tended to be affected more by the disease than women (Noymer and Garenne 2000). Male flu victims represented a population which in the absence of a pandemic would have been more at risk of contracting high-mortality endemic diseases, like tuberculosis (Noymer 2012).

Mortality statistics are widely used to track the severity of the Spanish flu and other pandemics. Because of the specific demographic factors which influence the course of any disease, academic discussions should, wherever possible, be making use of age-adjusted mortality rates based on vital statistics collected through civil registration. They should disaggregate their standardised mortality rates by sex, race and other categories of relevance. Direct comparisons within and between countries are difficult to make in the absence of such statistics. Any inferences drawn about the efficacy of public health policy, the speed of economic recovery or the electoral consequences of lockdown measures where the underlying demographic structures have not been accounted for cannot be robust. This is best illustrated with an example: if one location has a higher mortality rate than another, and also has a higher share of the population between 40-60 years of age, or a greater proportion of women, then we need to adjust for these differences because this location’s population is inherently less exposed to Influenza-18 than the other (i.e., it has a lower biological risk factor).^3^

Although we have long known that demography matters, the literature on Influenza-18 tends to rely on fragile population estimates, raw death counts and crude mortality rates. Most of last year’s new crop of pandemic economics studies make no acknowledgement of, let alone adjust for, the demographic composition of the countries and regions under study, either for 1918 or today (e.g., Lin and Meissner 2020). Even those studies which do take note of the differential demographic impact fail to consider its economic consequences.^4^ Many scholars even appear unaware that their chosen population statistics are arithmetic interpolations rather than real population counts (e.g., Correia et al. 2020). Studies that do incorporate population change fail to acknowledge the limitation of the underlying estimation methodology (e.g., Lilley et al. 2020). Beach et al. (2020), a new review of all aspects of the literature on the Spanish flu, largely overlooks issues stemming from demography. But a closer reading of some of the data sources used by researchers in this field would have highlighted the problem; contemporaneously it was acknowledged that ‘owing to recent unusual migrations of the population and the fact that 1916 [1917, 1918, 1919] is far away from the last census year, the [population] estimates are probably too high in some cases and too low in others’ (US CB 1918b, p. 62; 1919, p. 77; 1920, pp. 118-119; 1921, pp. 118-119).^5^

Why are economists ignoring demography? We think the answer lies with an apparent unawareness amongst our ranks of the standard statistics, and the data required to calculate these statistics, that demographers and health scientists use to understand mortality risks. We think this blind spot has led to scholars making use of “convenient” data, typically derived from the closest previous census. But this choice only amplifies the problem; we demonstrate that the use of census data distorts the denominator in mortality statistics – especially at a time of national crisis, such as during a war. We measure the consequence of this methodological choice using the case of Ireland. We show that choosing the convenient denominator means scholars fail to take full account of demographic change during World War I, a war in which large numbers of Irish men were sent to their deaths on the battlefields of Etreux, Gallipoli, Ypres, Hulluch, Passchendaele, the Somme and others.

Using what demographers call the “cohort component method”, which involves ‘a separate analysis of the changes affecting each component of the population’ (UN DESA 1956), we construct a new spatially disaggregated demographic dataset for the case of Ireland. Our analysis of this dataset suggests the very youngest in society, those under the age of five, had the highest mortality rates during the Spanish flu pandemic. Rather than focusing solely on young adults, our attention should therefore also be drawn to young children. Development and health economists have long pointed out that populations which survive catastrophic risks are selected populations with fundamentally different attributes to pre-crisis populations (Deaton 2007; see review in Blum et al. 2020). This insight may also apply to our case; the elimination of a cohort of young children may have fundamentally changed the attributes of Ireland’s surviving population into adulthood (cf. Almond 2005; Brown and Thomas 2018).

We think this finding has potentially significant implications for economic analyses linking mortality rates with other datasets, in this and other settings, historical or present day. Policymakers are currently making difficult choices about non-pharmaceutical public health interventions, designing targeted vaccination programmes, or weighing up the costs and benefits of financial support to firms, industries or regions. Our 100-year-old historical analogue demonstrates that carefully constructed population data and sensible demographic adjustments are necessary for our work to be useful to them. Health scientists are already starting to make such adjustments in their analyses of the Covid-19 pandemic (see, e.g., Bhopal and Bhopal 2020; Dowd et al. 2020; Kulu and Dorey 2020); economists must now follow suit.

## 3. The Irish Case

The impact of the Spanish flu on Ireland was first quantified by Ireland’s then-Registrar-General, Sir William John Thompson, in an article published immediately after the pandemic’s conclusion (Thompson 1919). Thompson limited his scope to influenza and pneumonia as the two causes of death associated with the pandemic.^6^ He estimated an influenza mortality rate of 243 per 100,000 in 1918, with urban areas experiencing a rate of 370 per 100,000. He ascribed to the Spanish flu 45 of the 140 per 100,000 who died of pneumonia. He calculated this excess mortality rate by comparing pneumonia deaths in 1918 with 1917. This brought Ireland’s total N1H1-related mortality rate to 288 per 100,000 for 1918.

The Irish experience of the Spanish flu was brought back to life again ten years ago in three PhD dissertations completed in close succession: by Caitríona Foley (examined at University College Dublin in 2009), Patricia Marsh (Queen’s University Belfast in 2010) and Ida Milne (Trinity College Dublin in 2011). Foley’s dissertation was subsequently published as a monograph, in 2011; Milne’s was published in 2018. Foley’s (2011) medical history-focused work revolves around how “ordinary people” reacted to the pandemic. She also recounts how Ireland’s medical professionals understood the science of infection and catalogues the various treatments they used. An exciting feature of her work is that she puts the Spanish flu into its long-run historical context, highlighting the fact that Ireland regularly suffered epidemics, including the so-called Russian flu in the early 1890s (Foley 2011, chap. 2).

Milne’s (2018) social history of the Spanish flu is a rich description of life during the pandemic, including details on the public policy response, particularly in the eastern province of Leinster. Her description of Ireland’s medical infrastructure highlights how the country’s local funding model for healthcare provisioning was incapable of dealing with a national health crisis. She describes how Irish officials had recently adopted international conventions on the collection and classification of health statistics, making Ireland’s official statistics comparable with other countries. However, as elsewhere in the world, she notes how doctors struggled to define cause of death; many individuals recorded as dying of tuberculosis, bronchitis, heart failure and other maladies probably died of the Spanish flu. Official statistics therefore likely underestimate the pandemic’s true impact, she argues. Milne estimates that 23,000 people died in the pandemic in Ireland, from which she infers one-fifth of the island’s population probably contracted the disease (Milne 2018, chap. 3). Using weekly data available only for the Dublin registry district, she calculates that the death rate for the city’s poorest social classes was probably almost double that of the richest (Milne 2018, chap. 3).^7^

Also very relevant to our research is Marsh’s (2010) unpublished dissertation on the Spanish flu in the northern province of Ulster. Of the three PhDs it includes the most extensive quantitative, demographic analysis. Marsh uncovers the precise timing of each of the three waves of the pandemic using a combination of official sources and local newspapers. Subsequently, she exploits the same official government sources we use in this current paper, and adopting the more sophisticated methodology of the then-Registrar General of England and Wales she re-calculate excess mortality statistics for Ireland using a broader list of causes of death than Thompson. She is therefore able to improve comparisons with the other two health statistics jurisdictions of the UK (England and Wales, and Scotland). Marsh estimates there may have been up to 14,000 additional deaths than previously ascribed to the flu, taking the total pandemic death toll to 34,000, and yielding a crude mortality rate of 782 per 100,000 population.^8^

The only work to make use of Irish influenza statistics in an econometric analysis is de Bromhead et al. (2020). The article is an analysis of the 1918 general election, which took place on 14 December – at the end of the second wave of the pandemic – and gave the previously-obscure Sinn Féin party the majority of Ireland’s 105 Westminster MPs. Using the same official sources we use, de Bromhead et al. calculate crude Influenza-18 mortality rates at the Poor Law Union level, the lowest administrative division at which health statistics were reported. They then use GIS software to allocate these to electoral constituencies – not an easy task since the two sets of boundaries do not line up. They find higher Spanish flu mortality is associated with a lower turnout on election day, but they argue this did not affect the overall electoral outcome.

To calculate the mortality rate, a demographic researcher requires a denominator: the relevant population, the average population exposed to risk of death during the defined time period. Up-to-date population estimates are necessary for a variety of administrative indicators, such as vital statistics and disease incidence (Long 1993). For their denominators, Thompson, Milne, Marsh and de Bromhead et al. all use the population taken from a census of Ireland conducted on 24 April 1911 (BPP 1913a). This choice is understandable as the most comprehensive sources for demographic data are censuses, and 1911 was the most recent census year.^9^

But populations can change, sometimes quite suddenly. Relying solely on the 1911 census means Thompson and the others do not take account of changes in population due to births, ageing, migration and deaths since 1911. Notably, the 1911 census fails to take account of the falling birth rates, the ageing population and changes in the composition of deaths due to warfare. The outdated denominator is compounded by a failure to age standardise Influenza-18 morality statistics.

Irish scholars of the Spanish flu are not alone in their choice to use the closest census. Around the world the most recent census year to the 1918-1919 pandemic were typically around 1910 (ex ante/pre-pandemic) or 1920 (ex post/post-pandemic), and so this same “error” is being repeated across the entirety of this literature. Age standardisation does not feature in any of last year’s new Influenza-18 studies.

## 4. Age Adjustment and Standardisation

To quantify the demographic impact of the Spanish flu, we need to estimate a denominator for our mortality statistics; relying on the closest pre- or post-pandemic census years will distort the true effects of the pandemic. At the most basic level, the population has changed in terms of births, ageing, migration and deaths in the intervening period. This was a very turbulent era, with the outbreak of a global war in August 1914 which lasted until November 1918, and a rebellion in Dublin City in April 1916; the 1911 census cannot take these into account. The pandemic was closely followed by a revolution, a guerrilla war, political partition of the island, large population resettlements of Protestants to Northern Ireland and a civil war in the newly-formed Irish Free State; the next censuses, conducted simultaneously in both Irish jurisdictions in 1926, is also not an appropriate choice for our denominator.

We need to make our own postcensal estimates of the population in 1918 and 1919 to calculate influenza-related mortality rates in 1918 and 1919:

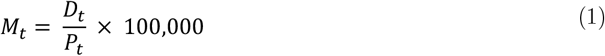

where *M*_*t*_ is the mortality rate per 100,000 population in year *t*; *D*_*t*_ is the number of deaths in year *t*; and *p*_*t*_ the base population in year *t*, typically measured mid-year. Because the nature of the disease means men and women were impacted differently, we need to calculate this separately by sex. After sex, age is the single most crucial variable when studying mortality (McGehee 2004); we need an accurate picture of the age structure of the population. Only once we have population by sex at each age, are we able to calculate age-specific mortality rates:

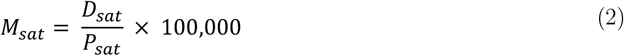

where *M*_*sat*_ is the mortality rate per 100,000 population for sex *s* at a specific age or age-range *a* in year *t*; *D*_*t*_ is the number of deaths for sex *s* at that age or age-range *a* in year *t*; and *p*_*sat*_ the base population for sex *s* in that age or age-range *a* in year *t*, typically measured mid-year. Alternatively, the crude death rate can be calculated as a weighted average of the death-by-age (*m*_*sat*_) and the population share of the relevant demographic (*p*_*n*_):

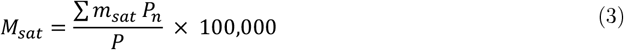

Demographers have long observed that comparisons of mortality using crude death rates alone can be misleading as the demographic (sex, age, race) composition of a population will affect the level of the observed death rate (see, e.g., Linder and Grove 1947, p. 60). Although the crude mortality rate is a weighted average mortality based on the age composition of a given population, we need to take account of age differences in order to make comparisons across populations with different age distributions. We can do this by imposing group weightings from a “standard” population. Doing this creates a hypothetical death rate which assumes the demographic composition of the population under study equals that of the standard population.

We report several estimates of a standard population. In order to make contemporary comparisons, we calculate a weighted average standard population for 1911 using data on OECD age structures that were reported in the 1911 *Census of England and Wales* (BPP 1917a, p. 63).^10^ We also compare these with four more modern standard populations: 1940 (US Standard Million), 1960 (World Standard Million), 2000 (WHO Standard Population), and 2011 (EU27 Standard) (NCI 2012; Ahmad et al. 2001).^11^ We report these standard population weights in <u>Table 1</u> for the relevant reporting age bands, alongside our own estimates for modern Ireland for comparison.^12^ The earlier age structures are more relevant for Influenza-18, when populations were younger on average; the latter standard populations are most relevant for Covid-19, where developed countries have populations that are older than developing countries on average.

**Table 1:** Distribution of the population in Ireland in 1911, 1918 and 1919, and standard population weights.

| Age (years) | Ireland |  |  |  |  |  | Standard Population Weights |  |  |  |  |  |
| --- | --- | --- | --- | --- | --- | --- | --- | --- | --- | --- | --- | --- |
|  | Male |  |  | Female |  |  | USA | World |  |  | EU27 | All-Ireland |
|  | 1911 | 1918 | 1919 | 1911 | 1918 | 1919 | 1940 | 1911 | 1960 | 2000 | 2011 | 2018 |
| 0 – 4 | 0.10 | 0.10 | 0.09 | 0.10 | 0.09 | 0.09 | 0.08 | 0.11 | 0.12 | 0.09 | 0.05 | 0.07 |
| 5 – 9 | 0.10 | 0.11 | 0.11 | 0.10 | 0.10 | 0.10 | 0.08 | 0.11 | 0.1 | 0.09 | 0.06 | 0.07 |
| 10 – 14 | 0.10 | 0.10 | 0.10 | 0.10 | 0.09 | 0.09 | 0.09 | 0.10 | 0.09 | 0.09 | 0.06 | 0.07 |
| 15 – 19 | 0.10 | 0.10 | 0.10 | 0.09 | 0.09 | 0.09 | 0.09 | 0.09 | 0.09 | 0.08 | 0.06 | 0.06 |
| 20 – 24 | 0.09 | 0.09 | 0.10 | 0.08 | 0.09 | 0.09 | 0.09 | 0.09 | 0.08 | 0.08 | 0.06 | 0.06 |
| 25 – 34 | 0.14 | 0.13 | 0.15 | 0.15 | 0.15 | 0.16 | 0.16 | 0.16 | 0.14 | 0.16 | 0.13 | 0.13 |
| 35 – 44 | 0.12 | 0.11 | 0.12 | 0.12 | 0.12 | 0.13 | 0.14 | 0.12 | 0.12 | 0.14 | 0.14 | 0.15 |
| 45 – 54 | 0.09 | 0.09 | 0.09 | 0.09 | 0.09 | 0.09 | 0.12 | 0.10 | 0.11 | 0.11 | 0.14 | 0.14 |
| 55 – 64 | 0.06 | 0.07 | 0.07 | 0.07 | 0.07 | 0.07 | 0.08 | 0.07 | 0.08 | 0.08 | 0.13 | 0.11 |
| Over 65 | 0.09 | 0.08 | 0.08 | 0.11 | 0.09 | 0.09 | 0.07 | 0.06 | 0.07 | 0.08 | 0.20 | 0.14 |

Source: 1911 census from the Census of Ireland conducted on April 1911 (BPP 1913a); 1918 and 1919 estimates are our calculation using method outlined in text with data from *Marriages, Births and Deaths Registered in Ireland* (BPP 1911-1919); source for standard population weights are NCI (2012) for USA, World and EU27, and CSO (2018) and NISRA (2019) for All-Ireland (Republic of Ireland plus Northern Ireland).

Here we adopt a direct age standardisation approach using:

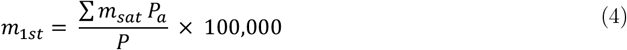

where *m*_*1st*_ refers to the mortality rate of the specific population under study; *p*_*a*_ is the standard population at each age; and *P* is the total standard population. Effectively this means we calculate a weighted average age-specific mortality rate.

In the existing Spanish flu literature there is a discrepancy over the term “age adjustment”. For example, Taubenberger (2006, p. 94) refers to ‘age-adjusted mortality data’, but the data that he refers to are actually age-specific mortality rates (death rates by different age bands); this is the same approach adopted by Milne (2018) for the case of Ireland. Age-specific mortality is useful for looking at a national picture. But demographic composition will distort spatial and temporal comparisons, so age standardisation is necessary; age adjustment needs to take account both the age-specific mortality *and* make an adjustment for differences in population structures by applying weights to age-specific mortality estimates.

## 5. Cohort Component Estimation

Ireland’s system of registration of births, marriages and deaths commenced in 1864.^13^Annual reports collated registration statistics from 130 reporting districts (Poor Law Unions) throughout the country and aggregated these to 32 counties, which are then further collapsed the island’s four historic provinces (Leinster, Munster, Ulster and Connacht). We rely on these data as our sources for *D*_*t*_ (the number of deaths in year *t*). The data were collated by age of death, sex, and cause of death. We digitised all *Registrar General Reports* between 1911 and 1920 to estimate our postcensal populations to use as the denominator in our mortality statistics (BPP 1912b, 1913c, 1914b, 1915b, 1916c, 1917c, 1918b, 1919b, 1920c, 1921b). The base population for our age structure estimates comes from the 1911 census (BPP 1913a), which was digitised by Clarkson et al. (1997).

To estimate the population in 1918 and 1919, we then use the cohort component method from demography (e.g. Bryan 2004; Watcheter 2014), which takes account of vital statistics in the intervening years (1911-1918). Essentially, we update the census with data on population flows. We use the following equation:

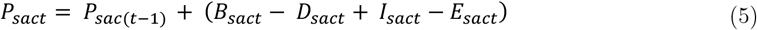

where *p*_*sat*_ is the population for sex *s* at age or age-range *a* in county *c* at time *t*; and *B, D, I* and *E* are within-year births, deaths, immigration and emigration, respectively.

The simple and cohort component methods of estimating population change have been considered the “gold standard” in population projection since at least World War II (UN DESA 1956), and they are still widely used by statistical agencies to estimate population (e.g., US CB 2014; ONS 2020).^14^ Population projections have their limitations and are not without controversy; demographers have missed post-war baby booms and subsequent baby lulls (De Gans 2002; Watcheter 2014). However, we do not suffer from this problem, as we use the method in a short contained timeframe to estimate historical populations and do not make assumptions about past trends.

We use annual records to help estimate postcensal populations between two decadal censuses at a spatially disaggregated level, along the lines described by Long (1996). National-level population estimates were estimated annually by contemporary government statisticians using a simple component method and were reported in the *Registrar General Reports*. Their methodology is straightforward: ‘by adding the births registered in each year to the estimated population for the previous year, deducting the deaths and the number of emigrants’ (BPP 1921b, p. 40). Essentially, we follow this same procedure at a district level for Ireland’s 32 counties, while also adjusting for ageing and migration.

We use direct measurement of births, deaths, and emigration. Contemporary immigration statistics are, unfortunately, harder to procure. We use information on annual immigration figures derived from Social Welfare (1955, p. 326).^15^ We focus here on international rather than internal migration.^16^ Deaths are recorded by age range in Ireland’s vital statistics, and so we can account for deaths within defined five-year age bins. Emigration age profiles are available from contemporary records (BPP 1912a, 1913b, 1914a, 1915a, 1916a, 1917b, 1918a, 1919a, 1920b). Immigration was considerably smaller than emigration in this period, a consequence of global warfare, so effectively this is a net migration story.

To estimate and annually update the age structure of the population, we move a share of each age bin forward one year, assuming a survival rate for the last year in the cohort. We explain the procedure for the age bin covering 25-29 year-olds as an example of our methodology. First, we account for all deaths and migration flows. Then, we bring a share of the population forward to the next bin; the 29-year-olds are moved to the 30-34 bin and are replaced in the 25-29 bin by 24-year-olds from the 20-24 bin. We adjust for the 24- and 29-year-old survival rates using weights calculated from the same age band in the 1926 census, the next census of Ireland following the 1911 census.^17^

We collapse all ages over 65 into one single age band as there are perceived discrepancies with age statements in the 1911 census, our base year. Ireland’s census commissioners believed the 1911 was more accurate than the previous census in terms of age statements as there was less heaping (reporting of rounded ages at 0 or 5) in the final report.^18^ The commissioners believed older people had in the past under-stated their age, but because of the recent introduction of the 1908 Old Age Pension for over-70s, they had for the first time ‘ascertained their correct age’ (BPP 1913a, p. 25). Ireland’s long history of youth emigration had left the country with an older population (Akenson 1993; Fitzpatrick 1980, 1984). It is likely that over-65s were therefore already a disproportionate share of the population, for demographic rather than nefarious reasons. However, Budd and Guinnane (1991) fear there was a deliberate overstatement of age to qualify for the pension. Collapsing the age bins over 65 years of age is our way to circumvent the possibility of overstatement of ages, while not trying to manipulate the underlying census.

Military enlistment during World War I is effectively treated as emigration in the Registrar General’s population estimates as it was a sizeable population movement to the battlefields of Europe. Subsequent demobilisation was treated as return migration (immigration). For example, in the population estimates for 1914, the male population decreases by 49,881, and in 1919 it increases by 63,000 (BPP 1921b). We follow a similar procedure. To adjust for military enlistment during World War I, we estimate the total enlistment in Ireland (134,202) from contemporary military sources (BPP 1921a, p. 9; WO 1922, p. 363).^19^

We detect the county composition of Irish enlistment from a further parliamentary source which covers 97 per cent of the total Irish recruits (BPP 1916b). The age of military service was between 19 and 41, but government statisticians noted ‘the male population of Ireland is composed chiefly of young men up to 18 years of age and of men over 50, as a large proportion of the remainder emigrates to the United States and Colonies’ (BPP 1921a, p. 9). Estimated military mortality is derived from War Office statistics, which imply a mortality rate of 14 per cent for all military personnel. Our war-related mortality estimate is lower than Bowman’s (2014) estimated 20 per cent mortality.^20^ We have chosen the more conservative death rate of 14 per cent to keep Irish mortality in line with UK-wide mortality figures.^21^

For cause-specific death rates in the pandemic, we report data on deaths attributed to influenza in addition to those attributed to all forms of pneumonia. While the inclusion of pneumonia with influenza deaths was common internationally, such as in the US (see US CB 1920, pp. 29-32; and used, e.g., in Brainerd and Siegler 2003), there are limitations of the data due to mistakenly attributing deaths to other causes, notably, to other respiratory illnesses (cf. Milne 2018, chap. 3). However, a no-pandemic counterfactual would also have seen many deaths due to influenza and pneumonia; despite being a great power, the UK remained a high-disease environment with a poor public health infrastructure, and so counting all who died of these two maladies would over-estimate the impact of Influenza-18. Equally, year-on-year fluctuations in the background mortality of certain age groups driven by other non-pandemic factors may result in an under-estimate of the pandemic’s true impact (Andreasen and Simonsen 2011).

To overcome these problems, we also report three excess mortality statistics: (1) deaths from *all* causes relative to what would “normally” have been expected across a given year; (2) deaths from influenza and pneumonia relative to normal; and (3) deaths from all causes for those aged under 45, the most severely-affected demographic group, relative to normal for that population group. Our estimates are based on a comparison of the mean influenza and pneumonia death rates for 1910-1914 with those in 1918 and 1919, calculated by sex and age. The selection of pre-war mean death rates is an attempt to not distort estimates with wartime spillovers. Moreover, 1915 saw a higher than usual number of deaths from tuberculosis, so including this year would distort the comparison. We report age-adjusted excess mortality using the estimated populations in 1918 and 1919 to account for changes in the composition of the population in the intervening period.

Our third measure of excess mortality follows an approach proposed by Andreasen and Simonsen (2011). The under-45s are well-documented across Influenza-18 studies to have suffered a disproportionate mortality burden. Consequently, Andreasen and Simonsen argue that including the over-45s in excess mortality calculations runs the risk of under-estimating the total burden of the pandemic because the background mortality fluctuations in older age groups may outweigh any impact of the pandemic. Excluding the over-45s means our excess mortality statistics are focused only on the population most at risk of pandemic-related death.^22^

Our full underlying cohort component population estimates for Ireland are available in a separate data paper (Colvin et al. 2021). Note that there exist various alternative methodologies for estimating population that are used to make population projections. One such alternative is linear interpolation. This method was used by US census officials to ensure timely estimates could be computed; ‘the method of arithmetical progression was adopted for computing the estimates of population […] based on the assumption that the increase each year since the enumeration is equal to the annual increase from 1900 to 1910’ (US CB 1918a, p. 5).^23^Adopting the interpolation methodology, given the data we now have available, would be rather crude and would hide variation within Ireland.^24^ Especially important in our case is that linear interpolation would yield misleading age compositions as it assumes trends from the previous decade are constant.^25^

## 6. Findings

We report our full county-level mortality statistics in the Appendix (<u>Tables A1 and A2</u>). Our starting point is a comparison of crude death rates across time, using <u>Table 2</u> and <u>Figure 1</u>. Ireland’s crude death rate increased from 16.52 per 1,000 in 1911, to a peak of 18.15 in 1918. Here we see a marked difference between Influenza-18 and Covid-19: the prevailing levels of mortality are higher as the Spanish flu pandemic occurs in an era of epidemiologic transition.^26^

**Table 2:**
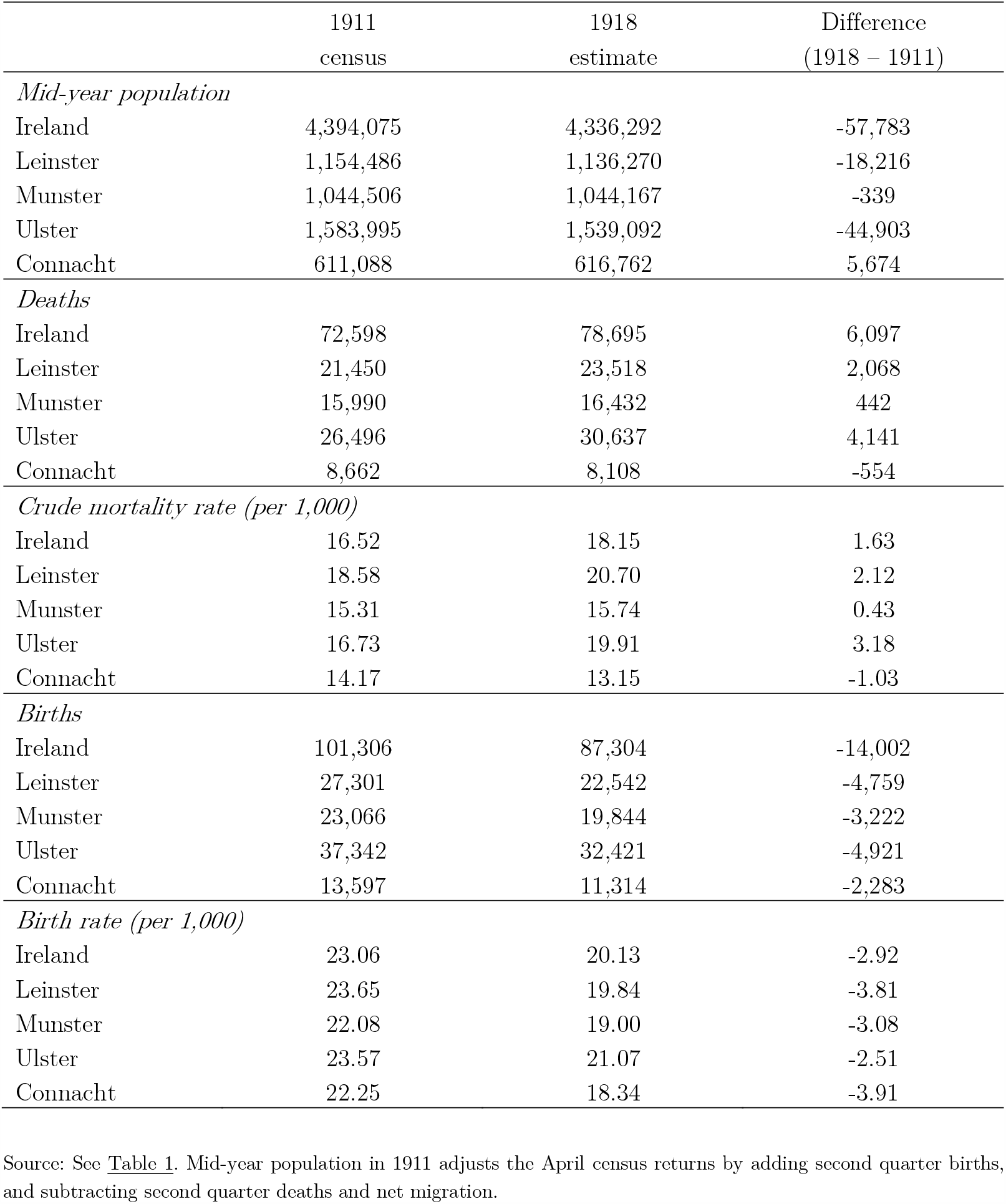
Demographic statistics for Ireland (1911 census versus 1918 estimate)

|  | 1911<br>census | 1918<br>estimate | Difference<br>(1918 – 1911) |
| --- | --- | --- | --- |
| *Mid-year population* |  |  |  |
| Ireland | 4,394,075 | 4,336,292 | -57,783 |
| Leinster | 1,154,486 | 1,136,270 | -18,216 |
| Munster | 1,044,506 | 1,044,167 | -339 |
| Ulster | 1,583,995 | 1,539,092 | -44,903 |
| Connacht | 611,088 | 616,762 | 5,674 |
| *Deaths* |  |  |  |
| Ireland | 72,598 | 78,695 | 6,097 |
| Leinster | 21,450 | 23,518 | 2,068 |
| Munster | 15,990 | 16,432 | 442 |
| Ulster | 26,496 | 30,637 | 4,141 |
| Connacht | 8,662 | 8,108 | -554 |
| *Crude mortality rate (per 1,000)* |  |  |  |
| Ireland | 16.52 | 18.15 | 1.63 |
| Leinster | 18.58 | 20.70 | 2.12 |
| Munster | 15.31 | 15.74 | 0.43 |
| Ulster | 16.73 | 19.91 | 3.18 |
| Connacht | 14.17 | 13.15 | -1.03 |
| *Births* |  |  |  |
| Ireland | 101,306 | 87,304 | -14,002 |
| Leinster | 27,301 | 22,542 | -4,759 |
| Munster | 23,066 | 19,844 | -3,222 |
| Ulster | 37,342 | 32,421 | -4,921 |
| Connacht | 13,597 | 11,314 | -2,283 |
| *Birth rate (per 1,000)* |  |  |  |
| Ireland | 23.06 | 20.13 | -2.92 |
| Leinster | 23.65 | 19.84 | -3.81 |
| Munster | 22.08 | 19.00 | -3.08 |
| Ulster | 23.57 | 21.07 | -2.51 |
| Connacht | 22.25 | 18.34 | -3.91 |

Source: See Table 1. Mid-year population in 1911 adjusts the April census returns by adding second quarter births, and subtracting second quarter deaths and net migration.

Another discernible aspect of <u>Figure 1</u> is the differences in the heights of the mortality spikes in 1918. One of the key distinctions between Ireland and these comparator countries is its unusual history of population decline from the 1850s onwards. The main driver of this decline was emigration; Ireland had the highest emigration rates in the world (Hatton and Williamson 1994, Tab. 1.1). These emigrants were young, and the residual population was therefore older. The inverse is true for immigrant-receiving countries, such as Australia or the US.

<u>Figure 2</u> makes use of quarterly returns of deaths in Ireland to estimate quarterly excess mortality by comparing quarters with average mortality in the same quarter of the previous four years. Here the striking aspect is the high excess mortality in the first quarter of 1900 almost rivals that of 1918. This spike is apparently the consequence of an influenza epidemic, where 4,677 deaths were attributed to influenza (199 per cent higher than the average of the previous four years) and 3,824 attributed to pneumonia (12 per cent higher than the average of the previous four years). The crude influenza mortality rate for 1900 was 106 per 100,000; age-adjusted, using the 1911 standard population in <u>Table 1</u>, the mortality rate was 99 per 100,000.^27^

Within Ireland, there are significant demographic differences in terms of migration and age structure. These are illustrated in the various population pyramids reported in <u>Figure 3</u>. Notably, urban centres in the east of the island grew, drawing internal migrants, while the countryside was depleted of youth. Comparing countries by age distribution – as done by contemporary census officials in BPP (1917a) – Ireland has one of the older populations; the share of over-55s was 16 per cent in Ireland versus an average of 12 per cent across other countries listed.^28^

**Figure 3:**
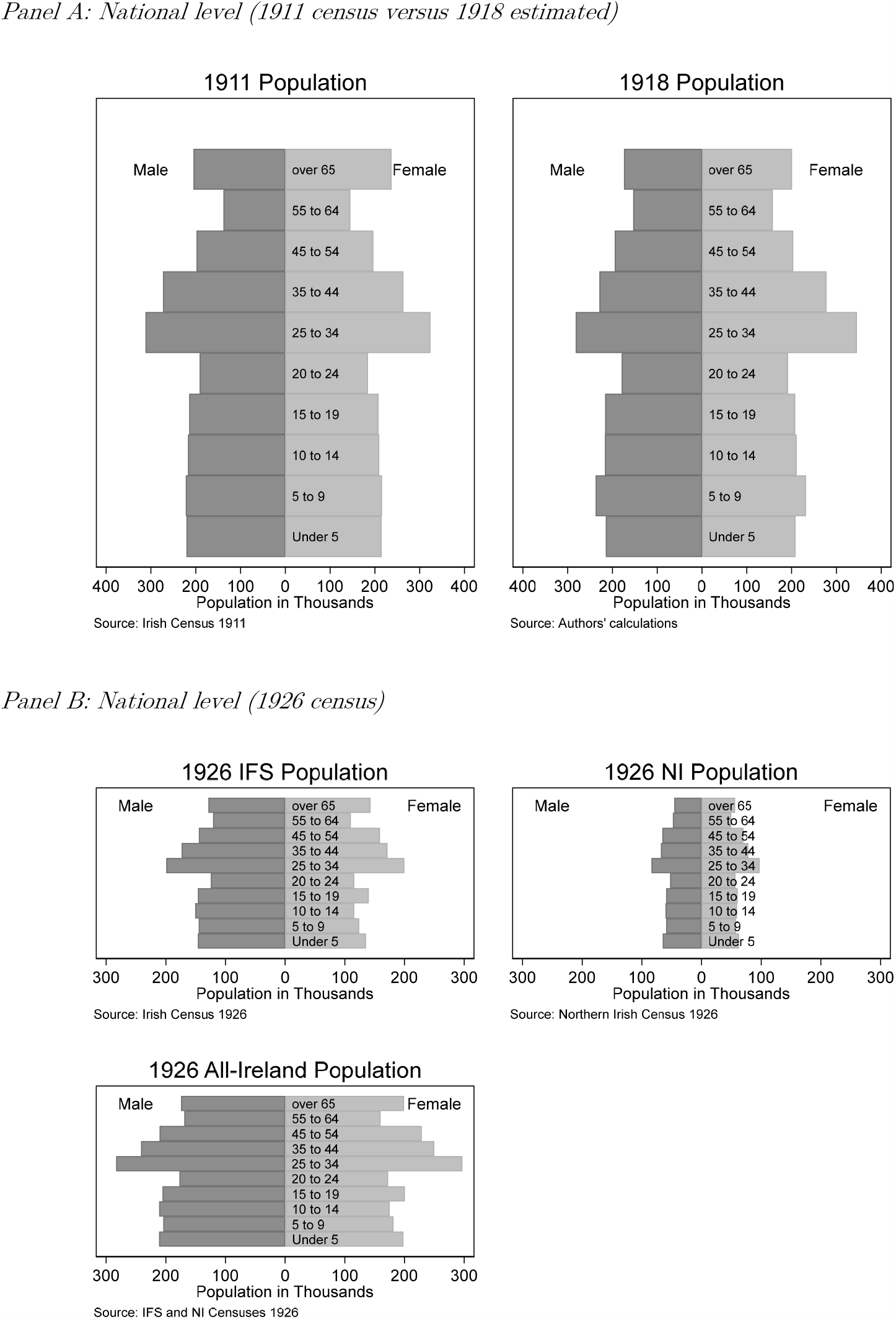
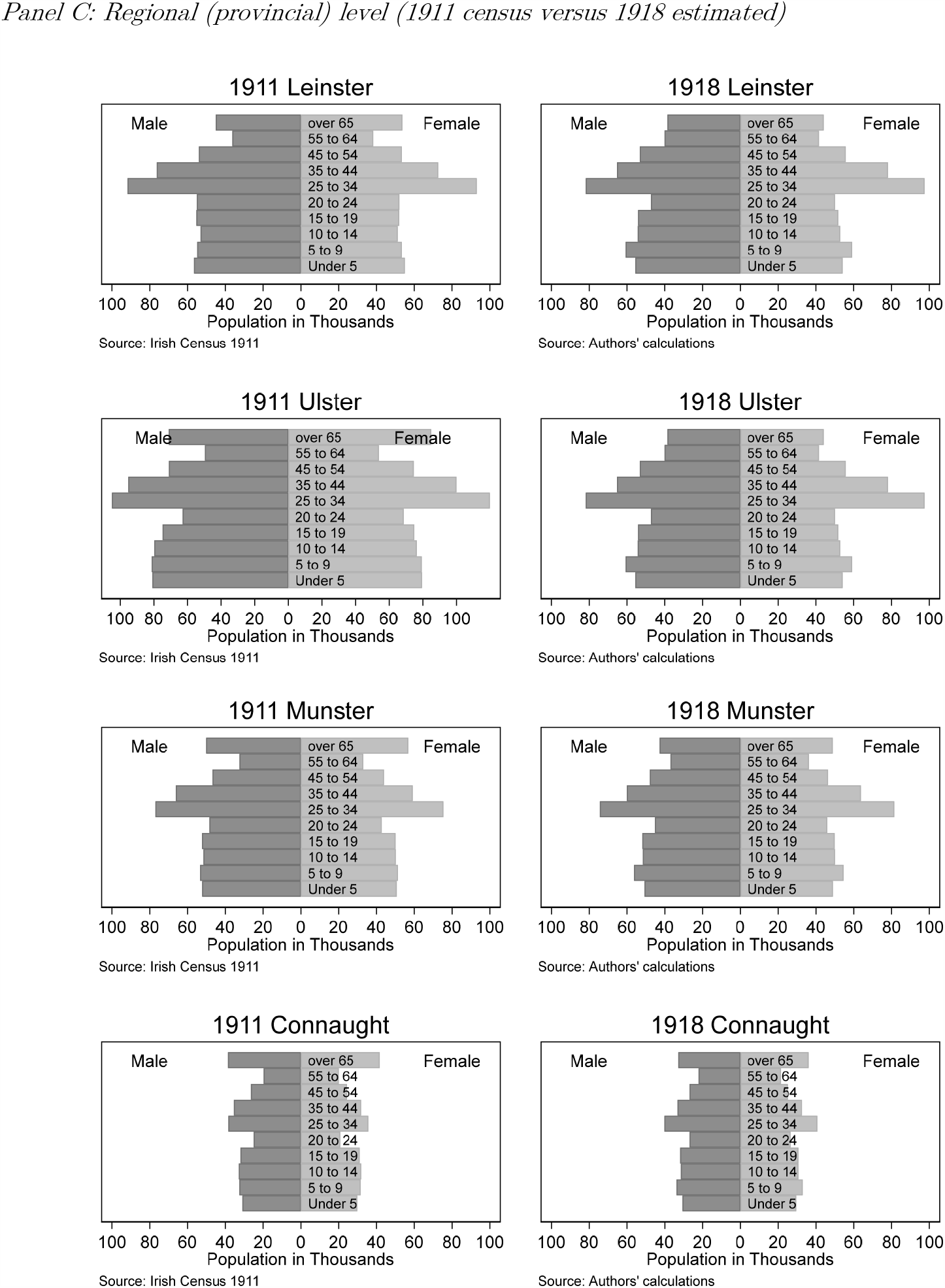
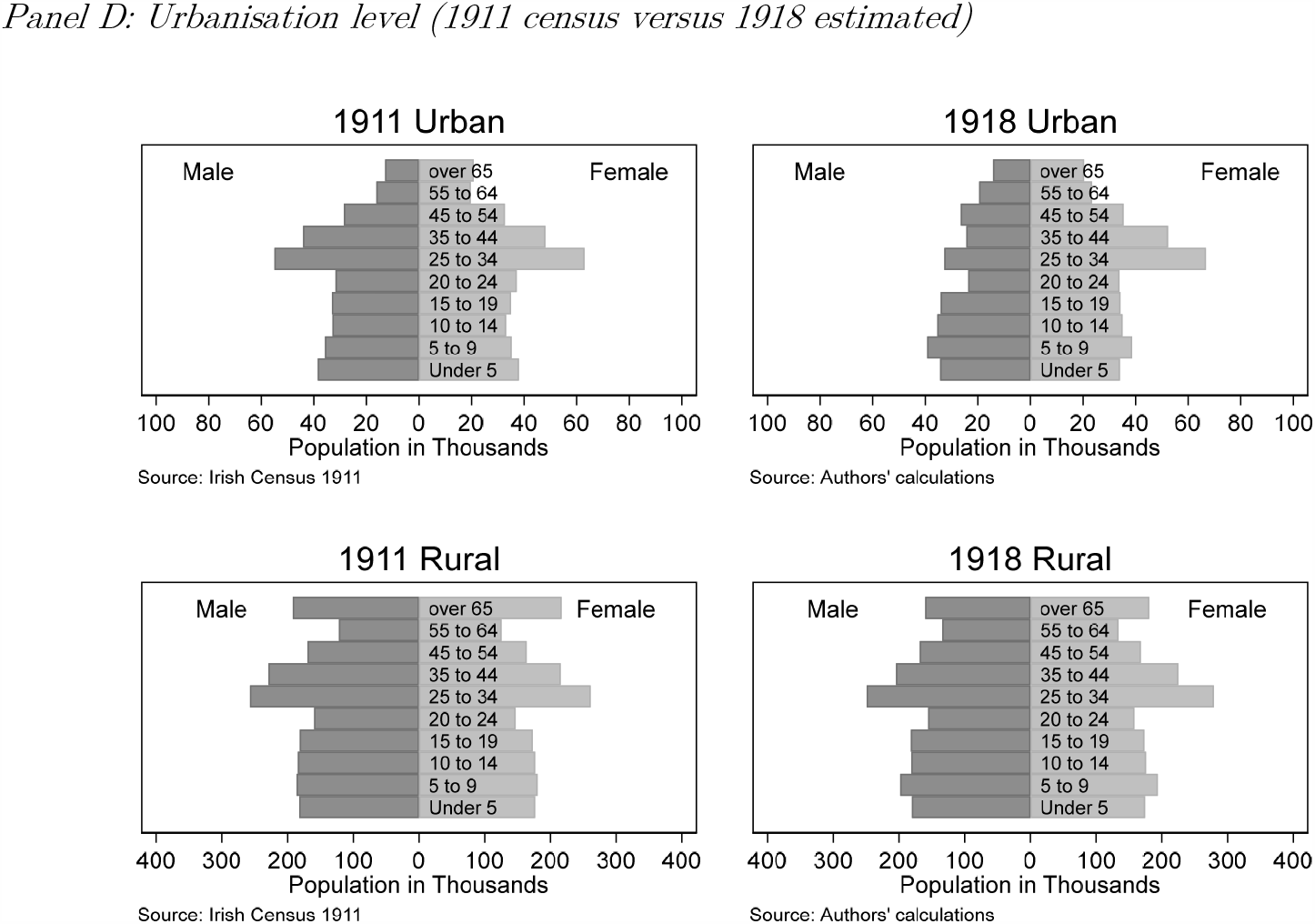
Population pyramids for Ireland. Note: Urban is defined here as Dublin City and Belfast, the island’s two main urban centres.

The population of Ireland decreased by 1.32 per cent in the seven years from the 1911 census (see <u>Table 2</u>). This fall in population was driven by a number of factors: a significant fall in birth rates, increasing mortality driven by an outbreak of pandemic tuberculosis in 1915, falling migration during the War, and military enlistment. If we ignore these demographic changes and use the 1911 population as our denominator for 1918 – the methodological choice made by Thompson (1919) and the others – then this implicitly, and incorrectly, assumes no change in the demographic composition of the population.

The changes in the composition are best illustrated through a visualisation of population pyramids. Firstly, as an aggregated national picture, <u>Figure 3</u> illustrates the compositional changes wrought by this declining population. There were noticeable falls in all age cohorts except the 5-9 and 55-64 cohorts (<u>Panel A</u> and <u>Panel B</u>). There is a distinct difference between male and female population change, as males made up the bulk of military recruits. Disaggregating to Ireland’s four historic provinces, we begin to see distinct regional patterns distinguishing the east and west of the island (<u>Panel C</u>). Further focusing in on the population in urban versus rural districts illustrates a disproportionate impact of military enlistment on cities (<u>Panel D</u>).

The raw data on influenza and pneumonia deaths in 1918 and 1919 are reported in <u>Table 3</u>. If we only look at the age distribution of deaths, we can confirm the “W-shaped” impact of the pandemic, where those aged 20-40 were disproportionately affected by Influenza-18: approximately 30 per cent of deaths hailed from people in this age group. However, this does not make allowance for the demographic weighting of these groups. <u>Table 4</u> calculates cause-specific death rates using both the 1911 census weights (<u>Panel A</u>) and our 1918/1919 estimated weights (<u>Panel B</u>). Across the board, the 1911 weights understate the impact of the pandemic. Adjusting for demographic changes, the Influenza-18 story is more nuanced, and, if anything, it aligns more with conventional mortality statistics, where the youngest and oldest display highest levels of mortality. This pattern is more clearly evident by comparing <u>Panel A</u> with <u>Panel B</u> of <u>Figure 4</u>.

**Table 3:** Raw influenza-related death counts, by age and sex (1918 and 1919)

| Age (years) | 1918 |  |  |  |  |  | 1919 |  |  |  |  |  |
| --- | --- | --- | --- | --- | --- | --- | --- | --- | --- | --- | --- | --- |
|  | Influenza |  |  | Influenza and pneumonia |  |  | Influenza |  |  | Influenza and pneumonia |  |  |
|  | Male | Female | *Total* | Male | Female | *Total* | Male | Female | *Total* | Male | Female | *Total* |
| 0 – 4 | 615 | 621 | *1,236* | 1,585 | 1,416 | *3,001* | 653 | 624 | *1,277* | 1,468 | 1,320 | *2,788* |
| 5 – 9 | 224 | 300 | *524* | 349 | 443 | *792* | 166 | 215 | *381* | 250 | 338 | *588* |
| 10 – 14 | 207 | 233 | *440* | 283 | 326 | *609* | 136 | 190 | *326* | 186 | 259 | *445* |
| 15 – 19 | 524 | 441 | *965* | 685 | 573 | *1,258* | 398 | 362 | *760* | 492 | 444 | *936* |
| 20 – 24 | 681 | 556 | *1,237* | 912 | 743 | *1,655* | 545 | 388 | *933* | 684 | 508 | *1,192* |
| 25 – 34 | 1,390 | 1,032 | *2,422* | 1,866 | 1,351 | *3,217* | 1,008 | 775 | *1,783* | 1,314 | 1,001 | *2,315* |
| 35 – 44 | 706 | 579 | *1,285* | 1,005 | 810 | *1,815* | 628 | 517 | *1,145* | 929 | 719 | *1,648* |
| 45 – 54 | 448 | 427 | *875* | 765 | 620 | *1,385* | 473 | 419 | *892* | 781 | 605 | *1,386* |
| 55 – 64 | 317 | 321 | *638* | 610 | 521 | *1,131* | 363 | 379 | *742* | 667 | 584 | *1,251* |
| Over 65 | 469 | 550 | *1,019* | 924 | 971 | *1,895* | 551 | 616 | *1,167* | 1,067 | 1,056 | *2,123* |
| Total | 5,581 | 5,060 | *10,641* | 8,984 | 7,774 | *16,758* | 4,921 | 4,485 | *9,406* | 7,838 | 6,834 | *14,672* |

Source: See Table 1.

**Table 4:** Influenza and pneumonia mortality rates, by age and sex (using both 1911 census and 1918/1919 estimated weights)

*Panel A: Mortality rates per 100,000 population (1911 census weights)*
| Age (years) | 1918 |  |  |  |  |  | 1919 |  |  |  |  |  |
| --- | --- | --- | --- | --- | --- | --- | --- | --- | --- | --- | --- | --- |
|  | Influenza |  |  | Influenza and pneumonia |  |  | Influenza |  |  | Influenza and pneumonia |  |  |
|  | Male | Female | *Total* | Male | Female | *Total* | Male | Female | *Total* | Male | Female | *Total* |
| 0 – 4 | 279 | 289 | *284* | 718 | 658 | *689* | 296 | 290 | *293* | 665 | 614 | *640* |
| 5 – 9 | 101 | 139 | *120* | 157 | 205 | *181* | 75 | 100 | *87* | 113 | 157 | *134* |
| 10 – 14 | 95 | 111 | *103* | 130 | 155 | *143* | 63 | 91 | *76* | 86 | 123 | *104* |
| 15 – 19 | 244 | 212 | *228* | 319 | 275 | *298* | 186 | 174 | *180* | 229 | 213 | *221* |
| 20 – 24 | 356 | 302 | *329* | 477 | 403 | *440* | 285 | 210 | *248* | 357 | 276 | *317* |
| 25 – 34 | 445 | 319 | *381* | 598 | 417 | *506* | 323 | 239 | *280* | 421 | 309 | *364* |
| 35 – 44 | 258 | 220 | *239* | 368 | 307 | *338* | 230 | 196 | *213* | 340 | 273 | *307* |
| 45 – 54 | 226 | 217 | *222* | 386 | 316 | *351* | 239 | 213 | *226* | 394 | 308 | *351* |
| 55 – 64 | 230 | 221 | *225* | 442 | 359 | *399* | 263 | 261 | *262* | 483 | 402 | *442* |
| Over 65 | 229 | 232 | *230* | 451 | 409 | *429* | 269 | 260 | *264* | 521 | 445 | *480* |
| Crude rate | 255 | 230 | *242* | 410 | 353 | *382* | 224 | 204 | *214* | 357 | 311 | *334* |
| *Age-standardised mortality rates* |  |  |  |  |  |  |  |  |  |  |  |  |
| 1911 World | 257 | 231 | *244* | 414 | 354 | *383* | 225 | 201 | *213* | 356 | 306 | *331* |
| 1940 USA | 262 | 232 | *247* | 414 | 349 | *381* | 230 | 204 | *217* | 359 | 304 | *332* |
| 1960 World | 255 | 231 | *243* | 418 | 358 | *387* | 227 | 205 | *216* | 366 | 315 | *340* |
| 2000 World | 260 | 232 | *246* | 416 | 352 | *384* | 230 | 206 | *218* | 365 | 310 | *337* |
| 2011 EU27 | 256 | 232 | *244* | 421 | 356 | *388* | 241 | 218 | *229* | 395 | 331 | *363* |
| 2018 All-Ireland | 255 | 230 | *242* | 416 | 352 | *384* | 235 | 212 | *223* | 381 | 322 | *351* |

*Panel B: Mortality rates per 100,000 estimated population (1918/1919 estimated weights)*
| Age (years) | 1918 |  |  |  |  |  | 1919 |  |  |  |  |  |
| --- | --- | --- | --- | --- | --- | --- | --- | --- | --- | --- | --- | --- |
|  | Influenza |  |  | Influenza and pneumonia |  |  | Influenza |  |  | Influenza and pneumonia |  |  |
|  | Male | Female | *Total* | Male | Female | *Total* | Male | Female | *Total* | Male | Female | *Total* |
| 0 – 4 | 286 | 298 | *292* | 738 | 679 | *709* | 312 | 307 | *310* | 702 | 650 | *676* |
| 5 – 9 | 94 | 129 | *112* | 147 | 191 | *169* | 71 | 94 | *82* | 106 | 148 | *127* |
| 10 – 14 | 95 | 110 | *103* | 130 | 154 | *142* | 63 | 90 | *76* | 86 | 123 | *104* |
| 15 – 19 | 242 | 212 | *228* | 317 | 276 | *297* | 184 | 173 | *178* | 227 | 212 | *220* |
| 20 – 24 | 380 | 290 | *333* | 509 | 387 | *446* | 256 | 200 | *229* | 321 | 261 | *293* |
| 25 – 34 | 493 | 298 | *386* | 662 | 391 | *512* | 302 | 222 | *261* | 393 | 286 | *339* |
| 35 – 44 | 308 | 208 | *254* | 439 | 292 | *358* | 234 | 185 | *209* | 346 | 257 | *300* |
| 45 – 54 | 230 | 210 | *220* | 392 | 304 | *347* | 246 | 205 | *225* | 406 | 296 | *349* |
| 55 – 64 | 206 | 203 | *205* | 397 | 330 | *363* | 234 | 238 | *236* | 431 | 366 | *398* |
| Over 65 | 269 | 274 | *272* | 530 | 483 | *505* | 324 | 315 | *319* | 626 | 540 | *580* |
| Crude rate | 266 | 226 | *245* | 428 | 347 | *386* | 223 | 201 | *211* | 702 | 306 | *330* |
| *Age-standardised mortality rates* |  |  |  |  |  |  |  |  |  |  |  |  |
| 1911 World | 274 | 225 | *248* | 438 | 348 | *390* | 222 | 198 | *210* | 356 | 304 | *329* |
| 1940 USA | 280 | 227 | *251* | 440 | 343 | *388* | 227 | 200 | *214* | 359 | 301 | *330* |
| 1960 World | 271 | 226 | *247* | 442 | 354 | *394* | 226 | 203 | *214* | 368 | 314 | *341* |
| 2000 World | 278 | 227 | *250* | 442 | 348 | *391* | 229 | 203 | *216* | 366 | 309 | *337* |
| 2011 EU27 | 276 | 232 | *252* | 452 | 359 | *402* | 246 | 220 | *233* | 408 | 339 | *372* |
| 2018 All-Ireland | 274 | 228 | *249* | 444 | 352 | *395* | 237 | 212 | *224* | 389 | 325 | *356* |

Source: See Table 1.

**Figure 4:**
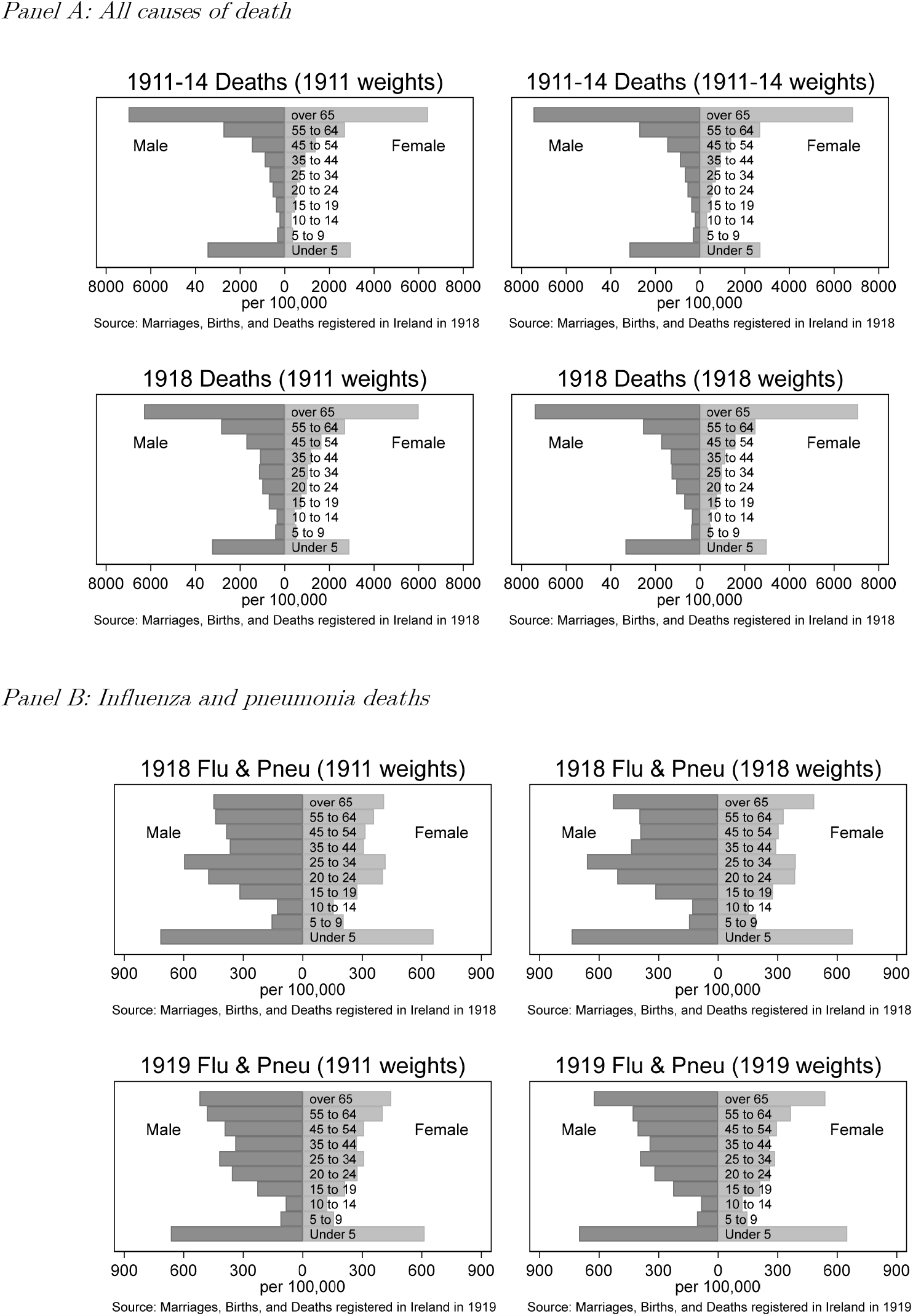
Mortality rates by age group (1911 census versus 1918 estimated weights)

<u>Table 5</u> reports distinct regional patterns in terms of age-specific and age-standardised mortality rates.^29^ Leinster and Ulster, located in the east of the island, show considerably higher age-standardised mortality than in Munster and Connacht, and are also high compared to the national average (see <u>Table 4, Panel B</u>). Focus on the main urban centres also shows much higher mortality across age groups, especially in the younger cohorts (under-5s), and age-standardised mortality rates in urban centres are more than double that of rural populations. This highlights the consequence of urban density for the spread of the disease. Disaggregated county maps of age-adjusted influenza-related mortality are presented in <u>Figure 5</u>, highlighting the considerable variation across the island.

**Table 5:** Influenza and pneumonia mortality rates per 100,000 in 1918, by region and urbanisation level (using 1918 estimated weights)

| Age (years) | Males |  |  |  |  |  | Females |  |  |  |  |  |
| --- | --- | --- | --- | --- | --- | --- | --- | --- | --- | --- | --- | --- |
|  | Leinster | Munster | Ulster | Connacht | *Urban* | *Rural* | Leinster | Munster | Ulster | Connacht | *Urban* | *Rural* |
| 0 – 4 | 922 | 488 | 962 | 251 | *1,901* | *517* | 892 | 447 | 846 | 240 | *1,674* | *484* |
| 5 – 9 | 178 | 130 | 163 | 80 | *250* | *127* | 260 | 139 | 221 | 76 | *370* | *155* |
| 10 – 14 | 184 | 68 | 160 | 67 | *243* | *109* | 233 | 110 | 173 | 46 | *257* | *134* |
| 15 – 19 | 379 | 204 | 417 | 150 | *501* | *282* | 345 | 184 | 347 | 130 | *516* | *228* |
| 20 – 24 | 589 | 422 | 602 | 304 | *828* | *460* | 485 | 248 | 498 | 154 | *723* | *315* |
| 25 – 34 | 900 | 441 | 787 | 318 | *1,424* | *561* | 481 | 228 | 502 | 152 | *650* | *329* |
| 35 – 44 | 633 | 318 | 492 | 165 | *1,087* | *362* | 360 | 202 | 343 | 138 | *493* | *245* |
| 45 – 54 | 496 | 257 | 455 | 275 | *751* | *336* | 344 | 201 | 403 | 108 | *547* | *253* |
| 55 – 64 | 461 | 324 | 460 | 244 | *718* | *350* | 358 | 229 | 420 | 200 | *588* | *285* |
| Over 65 | 674 | 376 | 702 | 246 | *954* | *492* | 535 | 315 | 696 | 225 | *695* | *459* |
| Crude rate | 556 | 304 | 514 | 208 | *848* | *362* | 429 | 228 | 441 | 147 | *639* | *289* |
| *Age-standardised mortality rates* |  |  |  |  |  |  |  |  |  |  |  |  |
| 1911 World | 564 | 308 | 532 | 211 | *917* | *365* | 434 | 229 | 440 | 143 | *661* | *285* |
| 1940 USA | 568 | 310 | 531 | 217 | *908* | *371* | 422 | 225 | 435 | 143 | *632* | *284* |
| 1960 World | 567 | 310 | 537 | 213 | *923* | *368* | 438 | 233 | 448 | 146 | *671* | *290* |
| 2000 World | 570 | 312 | 535 | 217 | *915* | *372* | 428 | 228 | 442 | 145 | *643* | *288* |
| 2011 EU27 | 582 | 321 | 550 | 227 | *911* | *389* | 428 | 236 | 468 | 157 | *626* | *308* |
| 2018 All-Ireland | 573 | 315 | 539 | 221 | *908* | *379* | 424 | 231 | 454 | 152 | *628* | *297* |

Source: See Table 1.

**Figure 5:**
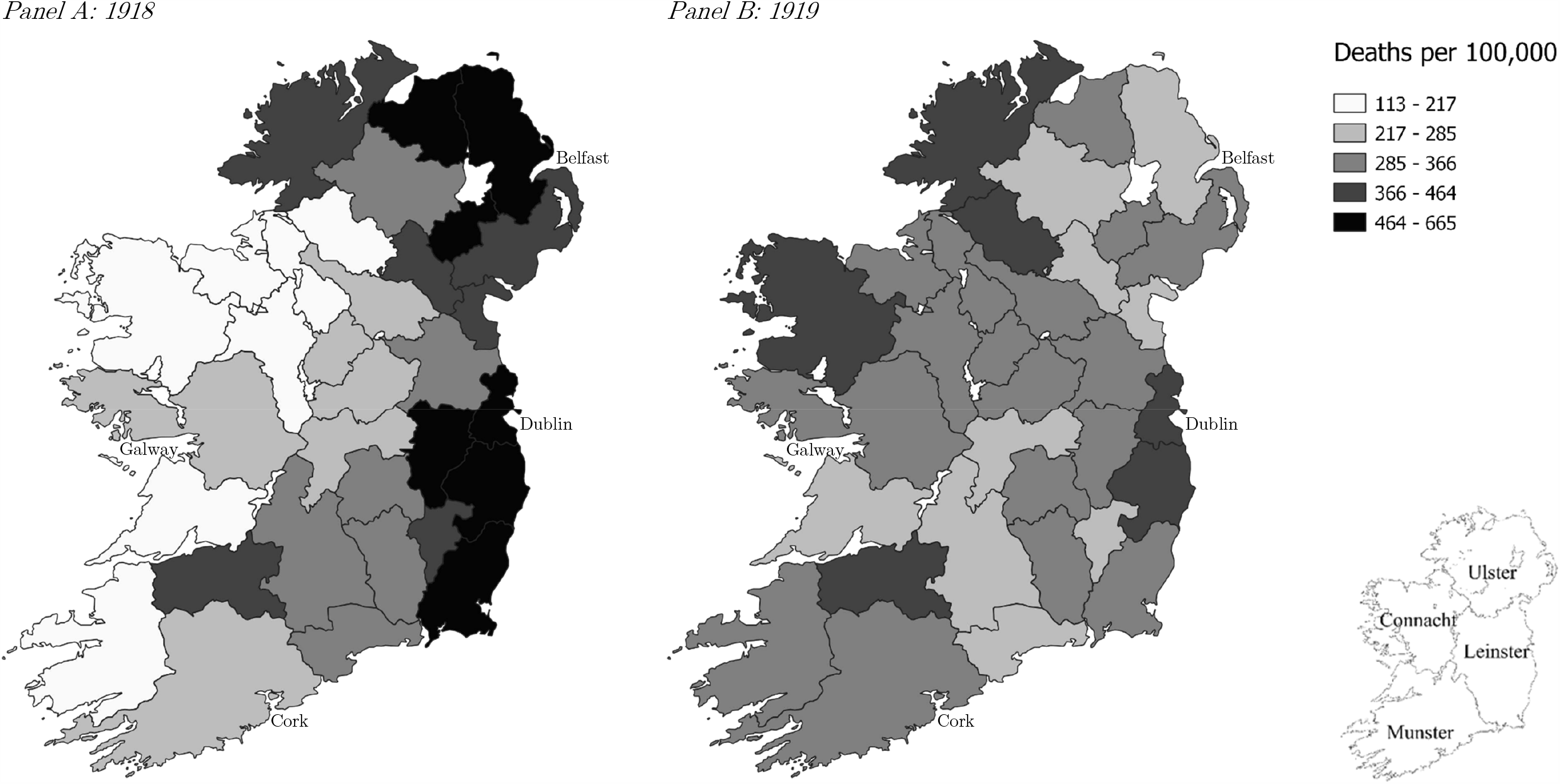
County map of age-adjusted influenza and pneumonia mortality rates (using 1918/1919 estimated weights) Note: Mortality rate for influenza and pneumonia. Underlying estimates reported in Appendix <u>Table A1</u>. Shading divided into five categories by equal quantiles for 1918. Dublin City is included in Dublin County; Belfast in Antrim.

Age-specific excess mortality, reported in <u>Figure 6</u>, show a similar pattern to age-specific influenza and pneumonia; with the exception that the oldest cohorts recorded fewer deaths in 1918 than they had recorded (on average) between 1911 and 1914, thus giving them negative excess mortality (see <u>Figure 7</u> for a map of age-adjusted excess mortality). Excess mortality more explicitly highlights the consequence of using the 1911 census rather than our estimated 1918/1919 denominators. The 1911 denominator records much lower than expected excess mortality in the oldest cohorts than the 1918 denominator.

**Figure 6:**
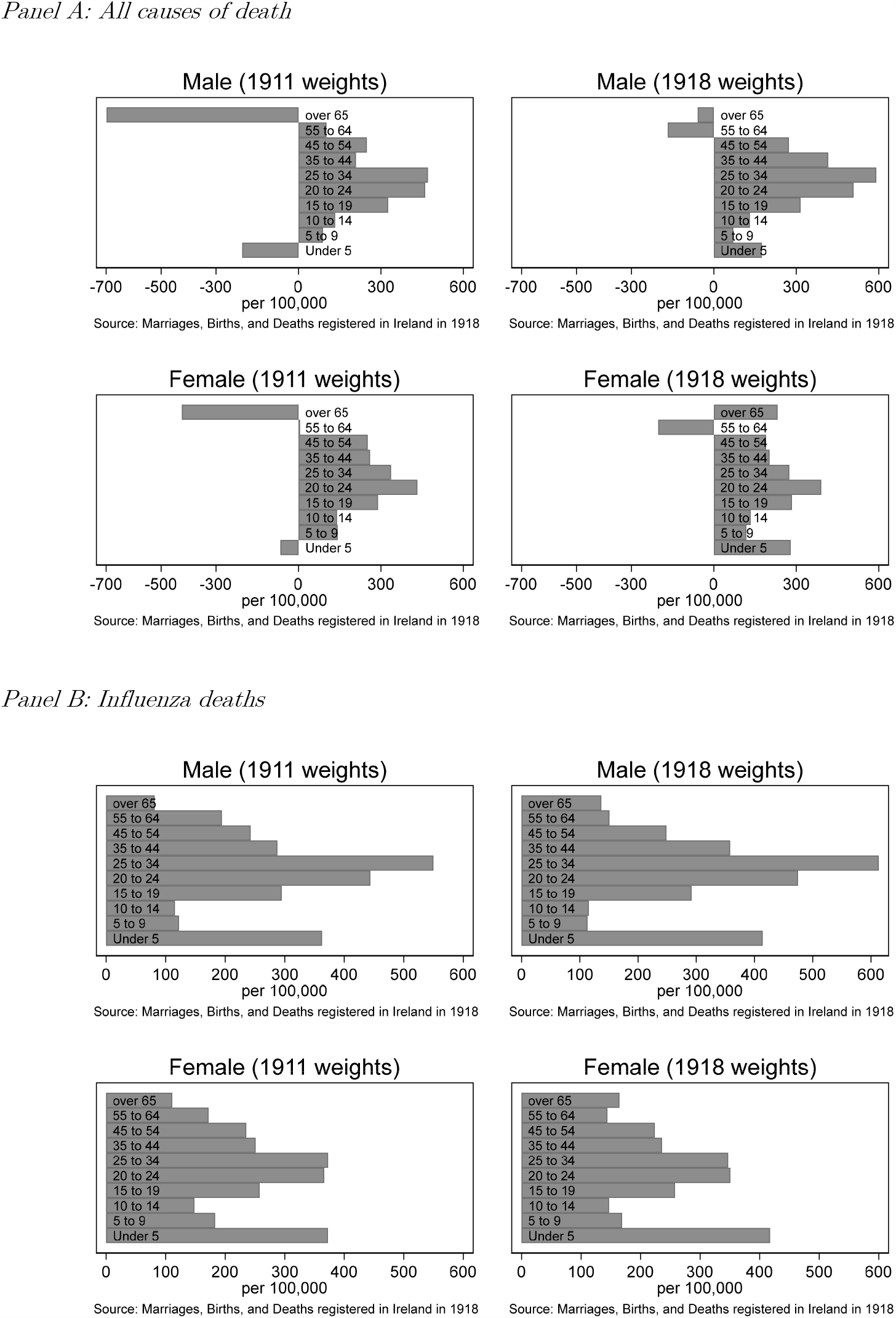
Excess mortality rates by age group in 1918 (compared to 1911-1914 average)

**Figure 7:**
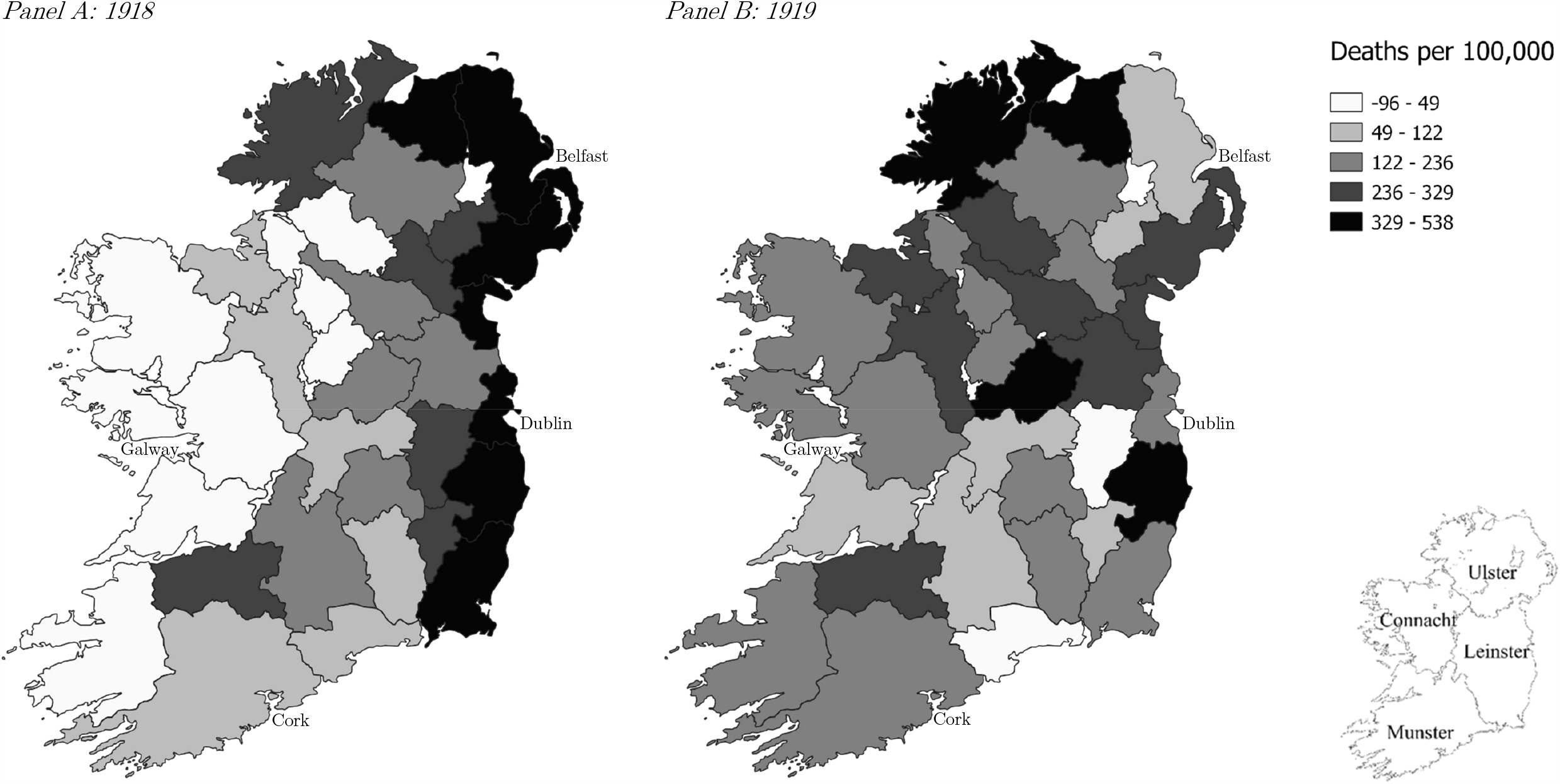
County map of age-adjusted excess mortality rates (compared to 1911-1914 average) Note: Underlying estimates reported in Appendix <u>Table A2</u>. Shading divided into five categories by equal quantiles for 1918. Dublin City is included in Dublin County; Belfast in Antrim.

In contrast to the cause-specific mortality figures shown in <u>Panel B of Figure 4</u>, the excess mortality figures of the prime working aged (25-34) is highest in <u>Panel B of Figure 6</u>. This figure, in turn, reflects the fact that the pandemic operated in a competitive disease environment. Indeed, comparing <u>Panel A with Panel B of Figure 6</u> suggests that whether the pandemic killed more children than usual is moot as there were many competing potential causes of childhood mortality in this era; similar numbers would probably have died in a no-pandemic counterfactual, but of other causes. Age-adjusted all-cause mortality shows a strong positive correlation with age-adjusted influenza and pneumonia mortality (<u>Figure 8</u>).

**Figure 8:**
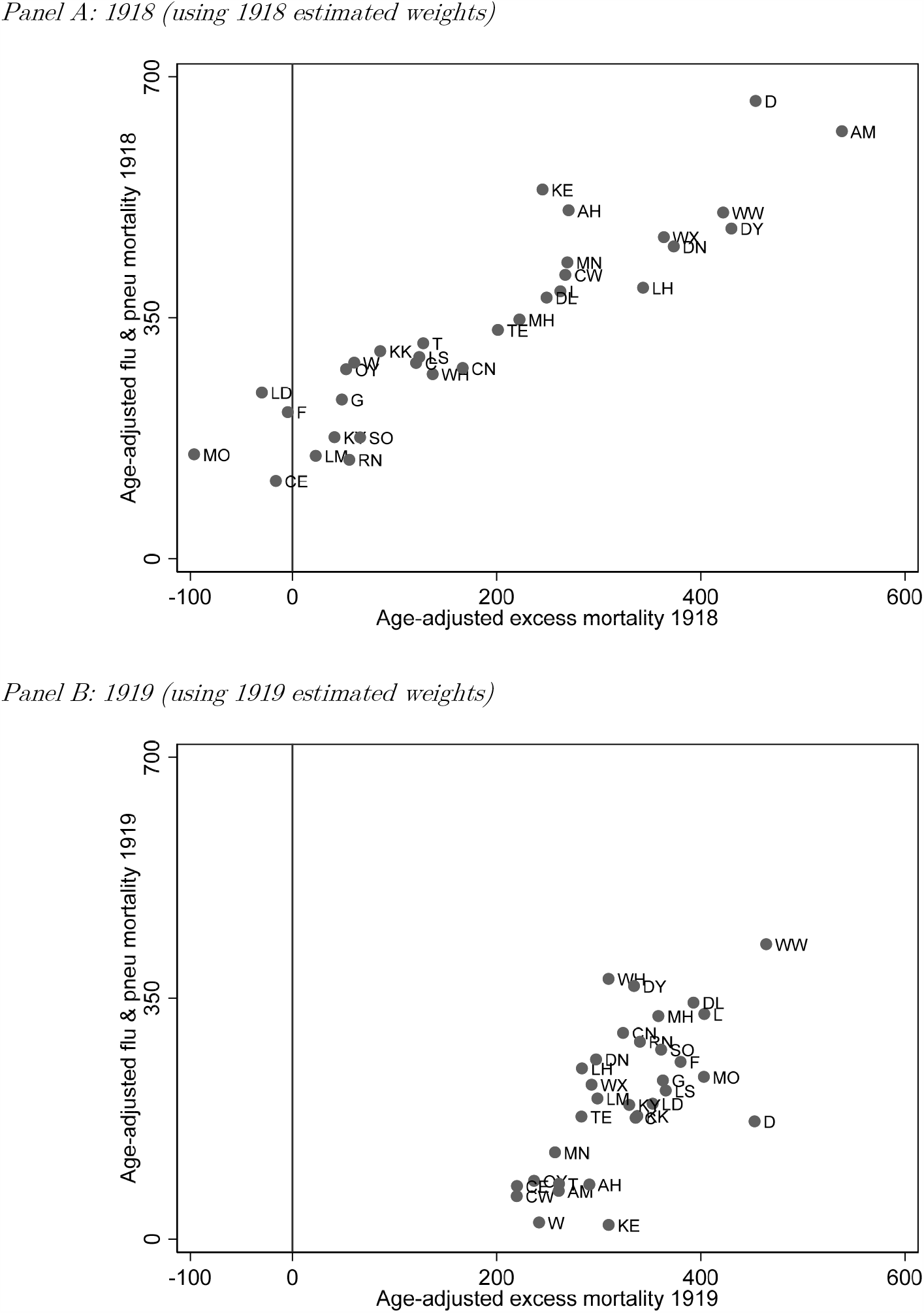
Scatter plots of age-adjusted mortality rates (influenza and pneumonia versus all-cause excess mortality) Note: Each point refers to a county or urban district using standard abbreviations. Underlying estimates reported in Appendix <u>Tables A1 and A2</u>. Source: Marriages, Births and Deaths registered in Ireland in 1918 and 1919.

Age of death-specific mortality rates suggest a possible mechanism of infection may have been family units (cf. Dowd et al. 2020a); across provinces, apart from Connacht, the ratio of the mortality rates of under-5s and 25-34s are of a similar magnitude.^30^ This trend is most pronounced for males as the female under-5 cohort has significantly higher mortality than the 25-34 cohort.^31^ This pattern suggests a possible confounder related to the economic activity of males, an idea raised independently by Marsh (2010) and Milne (2018). During World War I, part of Ireland’s labour force was deemed essential for both the war effort and for domestic morale. Reserved occupations in rural Ireland included farmers; the pastoral nature of the agricultural economy meant little social interaction. Reserved occupations in urban centres related to transport and factory work (railway and transport workers, food processing, shipbuilding and repairs), where what we would now call “social distancing” was near-impossible.^32^

The major puzzle is why those over 45 had lower than expected mortality (i.e., lower excess mortality compared to the 20-40 cohort). One explanation in the literature is they enjoyed some immunity thanks to previous exposure to influenza strains, such as that in 1900, which was particularly virulent (see Taubenberger 2006). These earlier influenza pandemics may also have selected the population in this age group, leaving only healthier individuals behind; the influenza death rate amongst the under-10s in 1900 was 64 per 100,000, compared with 25 and 80 per 100,000 for the 20-34 and 35-54-year-old cohorts. An alternative explanation is these older cohorts were less economically active and so less likely to catch influenza, or less likely to work while recovering from influenza (cf. Milne 2018).^33^

We have uncovered marked differences in demographic composition between 1911 and 1918. The most prominent divers of this change were war related. Most immediate was army enlistment from urban centres, which reduced the male population in the military-active age groups (20-40), the exact population identified in the epidemiology literature as being particularly susceptible to 1918’s N1H1 strain. Indirectly this led to a drop in birth rates during the war years. We also show rural centres were older owing to emigration trends – the specific population group traditionally identified as being less affected by the Spanish flu.

## 7. Discussion

As with the Great Recession referencing the Great Depression for influence in developing policy responses, the Great Virus of today sees continued reference to historical pandemics. The major analogue for Covid-19 is Influenza-18. The Spanish flu is now widely studied again and referred to both in academic and popular writing, including our own (Colvin and McLaughlin 2020a; Colvin 2020). The historical pandemic has notably been repeatedly referenced, albeit always incorrectly, by former US President Donald Trump (Rupar 2020; Mathers 2020). However, to fully extrapolate relevant policy lessons, we must first develop a more nuanced understanding of the population-at-risk in 1918. This means employing off-the-shelf methods from the demography literature to take account of the changing demographic composition of populations during the 1910s, a turbulent decade across the globe.

The existing literature on the 1918-1919 pandemic stresses the importance of age and sex, with males aged 20-40 constituting the population particularly at risk of succumbing to the virus. But surprisingly, the new economic studies of the pandemic mostly fail to address this fundamental issue of demography. They do not carry out age standardisation, without which mortality statistics across different localities are very difficult to compare. Economists must pay more attention to the source of their historical data, and take better note of the statistical methodologies developed by those in other disciplines to interpret these data. With the right data, age adjustment is a computationally straightforward procedure, and relevant standard populations are readily available.^34^

Additionally, the new Influenza-18 economics studies use unrepresentative population estimates in their mortality rate denominators. These have the potential to bias their results. The pandemic occurs late in the census cycle and starts during a world war which resulted in a global population upheaval. Using ex ante population estimates in mortality statistics implicitly assumes the prevailing decade had few demographic implications; adopting ex post population estimates overstates the burden of the pandemic as the dead are no longer counted in the population. Wherever possible, scholars should instead spend time to estimate their own denominators for their 1918 and 1919 mortality calculations, pieced together from readily available vital statistics published by national statistical agencies.

Quantitative demographic histories of Ireland end with World War I (see, e.g., Guinnane 1997) and therefore do not address demographic change across the specific period necessary to analyse the 1918-1919 pandemic. We must, therefore, carry out this analysis ourselves. Our new population estimates are reported in a separate paper (Colvin et al. 2021). Ireland’s experience of Influenza-18 has recently seen important contributions from social and medical historians (most notably: Marsh 2010 and Milne 2018). We complement their analysis by updating their figures with a more robust methodology which takes age-at-death into account. While we provide a more nuanced picture of the demographic impact of the flu, our total death toll attributed directly to influenza is not far off their overall estimates; we use the same sources after all.^35^

However, looking at excess mortality statistics tells a different story. Mean annual deaths were 72,706 between 1911 and 1914; 1918 saw 78,695 die, and in 1919 this was 78,612. Total all-cause excess deaths over both years were 11,895, considerably lower than the raw total influenza and pneumonia death counts. Meanwhile, excess influenza and pneumonia deaths (compared to similar deaths between 1911 and 1914) were 11,785 in 1918 and 2,829 in 1919. And all-cause excess deaths for those under the age of 45 were 7,292 in 1918 and 2,662 in 1919. The prevailing high mortality environment in which the pandemic occurred may account for this finding. Influenza crowded out other causes of death; people would likely have died from something else were instead killed by Influenza-18, a phenomenon known in epidemiology as the “harvesting effect” (Noymer 2012).

The all-cause excess mortality rate was 163 per 100,000 in 1918 and 115 per 100,000 in 1919. Adjusted for age, the statistics are 223 and 124 per 100,000, respectively. And focusing only on the under-45s, they are 240 and 62 per 100,000 respectively. Given the discrepancy in influenza-attributed mortality, excess mortality measures may provide a more grounded view of the impact of the 1918 pandemic. While this is something not readily discussed in the existing literature on Influenza-18 (e.g., Taubenberger 2006), those analysing Covid-19 have started to come around to this view (e.g., Beaney et al. 2020).

## 8. Influenza-18 and Covid-19

How do the methods employed to assess our historical case study help us to better understand our modern pandemic? To simplify our analysis, and to facilitate comparisons across time and space, we limit our study to the “Two Irelands”: the Republic of Ireland (the 26-county successor-state to the Irish Free State) and Northern Ireland (the six north-eastern counties of Ulster that remain part of the UK). We answer the question in a single figure: <u>Figure 9</u> reports population pyramids (<u>Panel A</u>) alongside Covid-19 mortality statistics for various age bands (<u>Panel B</u>) for the first four months of the pandemic (March to June 2020) in both Irish jurisdictions.^36^ The raw number of Covid-19 deaths in the Republic of Ireland for this period was 1,738; for Northern Ireland it was 837. Both polities show a similar distribution of Covid-19-attributed deaths by age: 93.5 per cent of attributed deaths in the Republic are over 65; for the North it is 92.8 per cent, a 0.7 percentage point difference.

**Figure 9:**
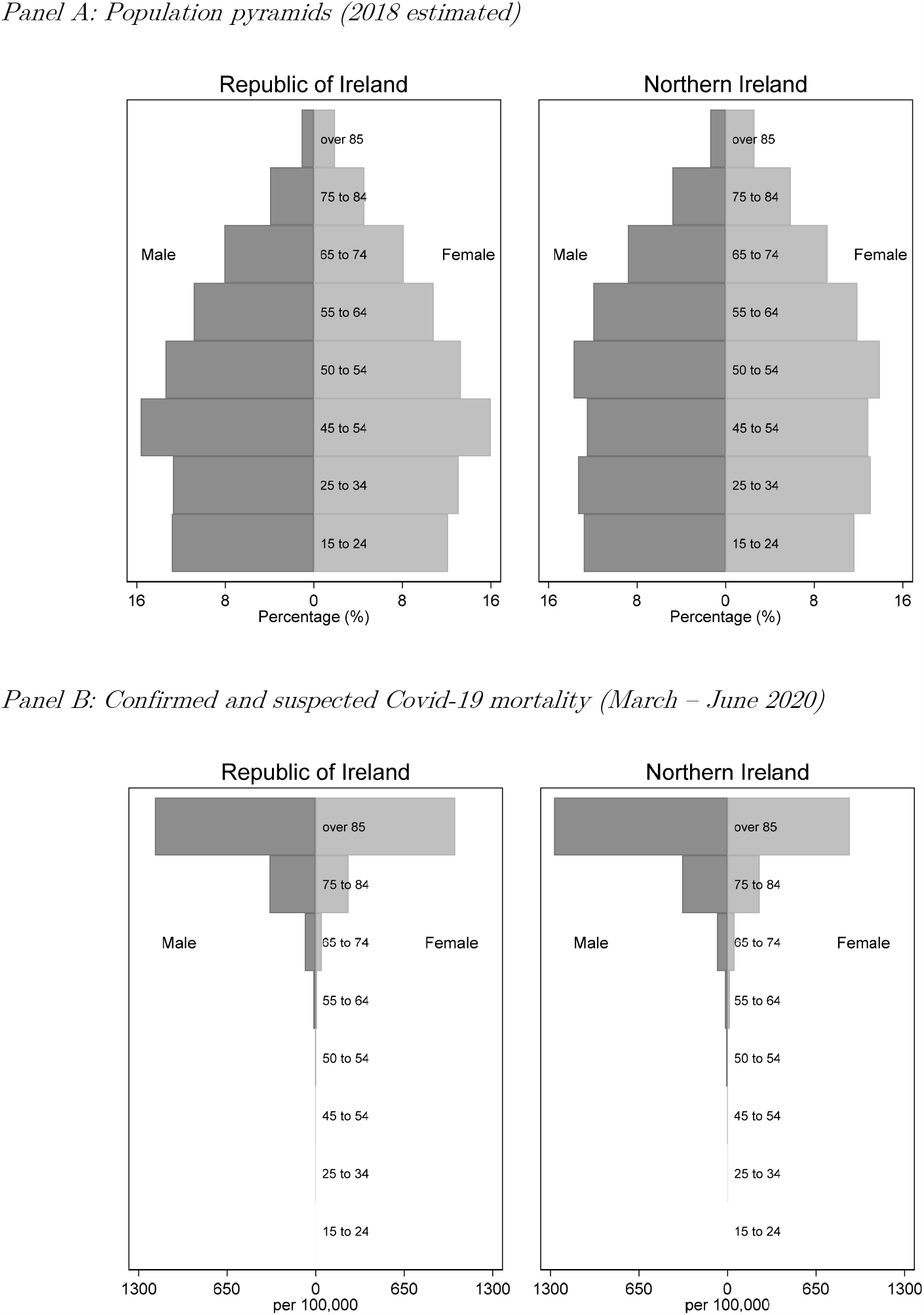
Population pyramids and Covid-19 mortality rates by age group. Note: Reporting period is 1 March to 28 June for the Republic of Ireland, and 1 March to 30 June for Northern Ireland. The CSO does not report exact numbers by sex below 5 out of privacy concerns. Data remain provisional, and some other unknown differences may exist in reporting standards. Source: For the Republic of Ireland: CSO (2018) and HPSC (2020a,b); for Northern Ireland: NISRA (2019, 2020).

To make a more meaningful comparison, we need to calculate mortality rates by age, and for this we need a denominator. We use the most recently available component method population estimates reported by each polity’s statistical agency, which pertain to mid-year 2018. There are notable age and sex composition differences between both Irish jurisdictions at the upper end of the age distribution, where the over-65 share is higher in the North (15.8 per cent, versus 13.9 per cent for the Republic).

We purposely have not annualised the Covid-19 death rates of these four months as we focus in only on the first wave of the pandemic. The raw Covid-19 mortality rate for the Republic of Ireland is 35.8 per 100,000 and for Northern Ireland is 44.5 per 100,000; the North’s rate is 24.3 per cent *higher* than the South’s. We then use the Republic’s age distribution as weights to estimate a standardised mortality rate for Northern Ireland: the North’s age-adjusted mortality rate is 35.6 per 100,000; after taking age into account, the North’s rate is 0.7 per cent *lower*. In the first phase of the Great Virus, our age-adjusted statistics suggest that differences in the mortality impact were negligible, despite the many policy differences on either side of the border.

Mortality differences by age reveal that in the 65-74 age band the North’s mortality rate was 2.7 per cent higher than the South, but in the 74-85 band the North was 2.4 per cent lower, while in the over-85s the North’s rate was 5.0 per cent lower. This point is not moot: deaths of over-85s accounted for 45.3 per cent of all Covid-19 attributed deaths in the South and 46.2 per cent of Covid-19 attributed deaths in the North. The age distributions of Covid-19 deaths were almost identical, but the underlying populations-at-risk is not. This point is entirely missed by media commentators.^37^ We therefore advocate that analysts take proper account of age in their statistical comparisons, and always present these alongside the crude mortality measures that have now become commonplace in policy discussions.

Of course, Covid-19 mortality rates remain tentative because of data availability issues and differences in reporting conventions. But they do point towards the idea that mortality rates which take account of demography can give new insights into where policy should be directed. In the case of the North, the disparity is in the mortality of the over-85s, particularly in men, where the mortality rate was 8.0 per cent higher than the rates of the same cohort in the South; for women the rate was 12.4 per cent lower than the rates in the South. A possible factor here could be the role of nursing homes: the North has 5.7 per cent more nursing home beds relative to the over-65 population than the South.^38^ The role of nursing homes in the current pandemic has proved particularly important and deserves special attention, in comparisons of the Two Irelands and further afield.^39^

In addition to enabling us to make comparisons across space in one pandemic, age adjustment also enables us to make comparisons across time at different pandemics. Thus, we can tentatively answer the question: how does Covid-19 compare with Influenza-18? We can do this using both historical and modern population standards from <u>Table 1</u>. However, here an issue becomes the choice of weights over time as Ireland’s 1918 population was lower and younger than that of the island today. Clearly, our historical pandemic was several orders of magnitude more serious in nature; using 2011 EU27 standard population weights, the Republic’s Covid-19 mortality is 65.0 per 100,000 and Northern Ireland is 64.1. These figures can be compared with age-adjusted mortality rates reported in <u>Tables 4 and 5</u>, where the adjusted mortality is approximately five times higher in 1918. This means we should probably caution drawing direct lessons about the impact of policy interventions from this historical pandemic.

It is important that economists take historical context into consideration in their comparisons. The 1918 outbreak occurred early in the epidemiological transition; deaths due to communicable diseases were more commonplace across what are today’s developed economies (<u>Figure 1</u>). Influenza-18 was a highly-communicable high-mortality disease which occurred in a high-disease environment, but apparently had few economic consequences (Velde 2020; Benmelech and Frydman 2020); Covid-19 is a highly-communicable low-mortality disease in a low-disease environment which has already incurred huge economic costs.

## Data Availability

Underlying data available from UK data archive

http://doi.org/10.5255/UKDA-SN-3578-1

## Appendix

**Table A1:** Crude and age-adjusted influenza-related mortality rates by county, 1918 and 1919.

| County | Crude mortality rates |  |  |  | Age-adjusted mortality rates |  |  |  | Difference |  |
| --- | --- | --- | --- | --- | --- | --- | --- | --- | --- | --- |
|  | Influenza and pneumonia |  | Difference with national crude |  | Age-adjusted (World 1911) |  | Difference with national crude |  | between crude and age-adjusted |  |
|  | per 100,000 |  | % |  | per 100,000 |  | % |  | percentage points |  |
|  | 1918 | 1919 | 1918 | 1919 | 1918 | 1919 | 1918 | 1919 | 1918 | 1919 |
| *Leinster* |  |  |  |  |  |  |  |  |  |  |
| Carlow | 404 | 225 | +4 | −27 | 412 | 220 | +6 | −33 | −1 | +6 |
| Dublin | 657 | 438 | +70 | +28 | 665 | 453 | +71 | +37 | 0 | −9 |
| Kildare | 544 | 303 | +41 | −7 | 536 | 310 | +38 | −6 | +3 | −1 |
| Kilkenny | 302 | 339 | −22 | +2 | 302 | 338 | −23 | +3 | +1 | 0 |
| King's* | 277 | 236 | −28 | −24 | 275 | 237 | −29 | −28 | +1 | +4 |
| Longford | 248 | 377 | −36 | +12 | 241 | 353 | −38 | +7 | +2 | +5 |
| Louth | 393 | 283 | +2 | −12 | 394 | 283 | +1 | −14 | +1 | +2 |
| Meath | 354 | 367 | −8 | +10 | 347 | 358 | −11 | +9 | +3 | +1 |
| Queen's** | 294 | 386 | −24 | +14 | 293 | 366 | −25 | +11 | +1 | +3 |
| Westmeath | 270 | 314 | −30 | −4 | 268 | 310 | −31 | −6 | +1 | +2 |
| Wexford | 458 | 291 | +18 | −10 | 467 | 293 | +20 | −11 | −1 | +1 |
| Wicklow | 498 | 461 | +29 | +34 | 503 | 464 | +29 | +41 | 0 | −7 |
| *Munster* |  |  |  |  |  |  |  |  |  |  |
| Clare | 109 | 210 | −72 | −31 | 113 | 220 | −71 | −33 | −1 | +2 |
| Cork | 283 | 335 | −27 | +1 | 284 | 336 | −27 | +2 | 0 | −1 |
| Kerry | 174 | 322 | −55 | −2 | 177 | 330 | −55 | 0 | 0 | −2 |
| Limerick | 380 | 402 | −2 | +19 | 388 | 403 | 0 | +22 | −1 | −4 |
| Tipperary | 319 | 260 | −17 | −18 | 313 | 261 | −20 | −21 | +2 | +3 |
| Waterford | 290 | 248 | −25 | −21 | 285 | 241 | −27 | −27 | +2 | +6 |
| *Ulster* |  |  |  |  |  |  |  |  |  |  |
| <u>Antrim</u> | 595 | 245 | +54 | −22 | 621 | 261 | +59 | −21 | −5 | −1 |
| <u>Armagh</u> | 492 | 300 | +27 | −8 | 506 | 291 | +30 | −12 | −3 | +4 |
| Cavan | 292 | 327 | −25 | −1 | 277 | 324 | −29 | −2 | +4 | +1 |
| Donegal | 404 | 415 | +5 | +22 | 379 | 393 | −3 | +19 | +7 | +3 |
| <u>Down</u> | 452 | 296 | +17 | −9 | 454 | 297 | +16 | −10 | +1 | +1 |
| <u>Fermanagh</u> | 226 | 393 | −42 | +16 | 213 | 380 | −45 | +15 | +4 | +1 |
| <u>Londonderry</u> | 486 | 345 | +26 | +4 | 480 | 334 | +23 | +2 | +3 | +2 |
| Monaghan | 436 | 262 | +13 | −18 | 430 | 257 | +10 | −22 | +2 | +4 |
| <u>Tyrone</u> | 345 | 303 | −11 | −7 | 332 | 283 | −15 | −14 | +4 | +7 |
| *Connacht* |  |  |  |  |  |  |  |  |  |  |
| Galway | 233 | 367 | −40 | +10 | 231 | 363 | −41 | +10 | +1 | −1 |
| Leitrim | 154 | 305 | −60 | −7 | 149 | 299 | −62 | −9 | +2 | +3 |
| Mayo | 150 | 394 | −61 | +17 | 152 | 403 | −61 | +22 | 0 | −6 |
| Roscommon | 146 | 337 | −62 | +2 | 144 | 340 | −63 | +3 | +1 | −1 |
| Sligo | 174 | 355 | −55 | +6 | 176 | 361 | −55 | +10 | 0 | −3 |
| *National* | *386* | *330* |  |  | *390* | *329* |  |  | *+1* | *+1* |
|  |  |  |  |  |  |  |  |  | *(SD 2)* | *(SD 4)* |

Note: \* King's County is now known as County Offaly. \*\* Queen's County is now County Laois. Underlined constitute the six counties of Northern Ireland from 1921.
Source: See Table 1.

**Table A2:** Crude and age-adjusted excess mortality rates by county, 1918 and 1919.

| County | Crude excess mortality rates |  |  |  | Age-adjusted excess mortality rates |  |  |  | Difference |  |
| --- | --- | --- | --- | --- | --- | --- | --- | --- | --- | --- |
|  | Excess |  | Difference with |  | Age-adjusted |  | Difference with |  | between crude |  |
|  | mortality |  | national crude |  | (1911 World) |  | national crude |  | and age-adjusted |  |
|  | 1918 | 1919 | 1918 | 1919 | 1918 | 1919 | 1918 | 1919 | 1918 | 1919 |
|  | per 100,000 |  | % |  | per 100,000 |  | % |  | percentage points |  |
| *Leinster* |  |  |  |  |  |  |  |  |  |  |
| Carlow | 212 | 1 | +31 | −99 | 267 | 62 | +20 | −50 | +11 | −49 |
| Dublin | 435 | 89 | +167 | −23 | 454 | 171 | +103 | +38 | +64 | −61 |
| Kildare | 195 | −56 | +20 | −149 | 245 | 21 | +10 | −83 | +10 | −65 |
| Kilkenny | −33 | 76 | −120 | −34 | 86 | 179 | −61 | +44 | −59 | −78 |
| King's* | −27 | 3 | −116 | −98 | 52 | 85 | −76 | −32 | −40 | −66 |
| Longford | −143 | 131 | −188 | +13 | −30 | 197 | −113 | +58 | −74 | −45 |
| Louth | 289 | 174 | +78 | +51 | 343 | 248 | +54 | +99 | +24 | −49 |
| Meath | 133 | 251 | −18 | +117 | 222 | 324 | 0 | +161 | −18 | −43 |
| Queen's** | −13 | 113 | −108 | −2 | 124 | 216 | −44 | +74 | −64 | −76 |
| Westmeath | 78 | 317 | −52 | +174 | 137 | 378 | −38 | +204 | −14 | −30 |
| Wexford | 259 | 133 | +59 | +16 | 364 | 224 | +63 | +80 | −4 | −65 |
| Wicklow | 309 | 273 | +90 | +136 | 422 | 428 | +89 | +245 | +1 | −108 |
| *Munster* |  |  |  |  |  |  |  |  |  |  |
| Clare | −112 | 42 | −169 | −64 | −16 | 77 | −107 | −38 | −61 | −26 |
| Cork | 49 | 111 | −70 | −4 | 121 | 176 | −46 | +42 | −24 | −45 |
| Kerry | −16 | 153 | −110 | +33 | 41 | 195 | −82 | +57 | −28 | −24 |
| Limerick | 143 | 221 | −12 | +91 | 262 | 327 | +18 | +163 | −30 | −72 |
| Tipperary | 55 | 8 | −66 | −93 | 128 | 80 | −43 | −35 | −24 | −58 |
| Waterford | 13 | −44 | −92 | −138 | 61 | 24 | −73 | −81 | −19 | −58 |
| *Ulster* |  |  |  |  |  |  |  |  |  |  |
| <u>Antrim</u> | 484 | −13 | +198 | −111 | 538 | 70 | +141 | −44 | +57 | −67 |
| <u>Armagh</u> | 151 | −34 | −7 | −130 | 270 | 79 | +21 | −36 | −28 | −93 |
| Cavan | 57 | 222 | −65 | +92 | 167 | 300 | −25 | +141 | −40 | −49 |
| Donegal | 159 | 301 | −2 | +160 | 249 | 344 | +12 | +176 | −14 | −16 |
| <u>Down</u> | 311 | 187 | +91 | +62 | 373 | 261 | +67 | +110 | +24 | −48 |
| <u>Fermanagh</u> | −139 | 114 | −186 | −1 | −5 | 257 | −102 | +107 | −84 | −108 |
| <u>Londonderry</u> | 348 | 293 | +114 | +154 | 430 | 368 | +93 | +196 | +21 | −42 |
| Monaghan | 178 | 31 | +10 | −73 | 269 | 126 | +21 | +1 | −11 | −75 |
| <u>Tyrone</u> | 113 | 84 | −30 | −27 | 201 | 178 | −10 | +43 | −21 | −70 |
| *Connacht* |  |  |  |  |  |  |  |  |  |  |
| Galway | −35 | 169 | −121 | +47 | 48 | 230 | −78 | +85 | −43 | −39 |
| Leitrim | −30 | 206 | −119 | +78 | 23 | 204 | −90 | +64 | −29 | +14 |
| Mayo | −184 | 180 | −213 | +56 | −96 | 236 | −143 | +90 | −70 | −34 |
| Roscommon | −28 | 241 | −117 | +109 | 56 | 287 | −75 | +131 | −42 | −22 |
| Sligo | 3 | 215 | −98 | +86 | 66 | 275 | −70 | +121 | −28 | −35 |
| *National* | *163* | *115* |  |  | *223* | *124* |  |  | *−20* | *−53* |
|  |  |  |  |  |  |  |  |  | *(SD 34)* | *(SD 26)* |

Note: See Table A1.
Source: See Table 1.

## Footnotes

1 There is significant disagreement in the literature on the global mortality burden of the Spanish flu, with popularly-cited estimates of between 24.7 and 39.3 million at one end of the spectrum (Patterson and Pyle 1991), and between 50 and 100 million at the other (Johnson and Mueller 2002). An indirect estimation of all-cause pandemic-related mortality by Spreeuwenberg et al. (2018) revises the death toll significantly downwards, to 17.4 million.

2 Note that differences in death certification standards between jurisdictions make international comparisons challenging (see Alderson 1981, pp. 34-40, for discussion of different registration systems across countries). This is true also today (see, e.g., Martin 2020).

3 A relevant modern comparison used in Dowd et al. (2020) is that of Brazil and Nigeria. In Brazil today 21% of the population is under the age of 15 and 10% over 65; in Nigeria 44% is under-15 and 3% is over 65. If a disease, like Covid-19, disproportionately affects the old and leaves the young unharmed, then comparative discussions of its impact must consider that Brazil’s population is inherently more vulnerable than Nigeria’s.

4 Correia et al. (2020), for example, exploit regional variation in Spanish flu mortality in the US to look at the pandemic’s subsequent economic impact, but do not control for regional heterogeneity in population structure.

5 After the 1920 US census, estimated growth by state was re-estimated as ‘the populations are estimated by the arithmetical method based on the 1910 and 1920 censuses’ (US CB 1922, p.74).

6 Ireland’s Registrar-General adopted a more straightforward methodology than his contemporary counterparts in London for their quantification of the mortality burden in England and Wales, who also included bronchitis, heart disease and phthisis (BPP 1920a).

7 Elsewhere, Milne (2019) describes how she collected oral histories of the pandemic, which she argues add an understanding which ‘cannot be discovered or quantified by the Registrar-General’s statistics’ (p. 16).

8 Elsewhere, Marsh (2019) further discusses some of her PhD’s findings on gender and mortality in the case of Ulster. She argues Ulster’s higher female fatalities may have been driven by the province’s higher female-to-male ratio.

9 This census was based on a house-to-house collection of data and was considered by census officials to be their best-yet.

10 For OECD countries, we take: Australia, Austria, Belgium, Canada, Denmark, England and Wales, France, Germany, Ireland, Italy, the Netherlands, Norway, Portugal, Scotland, South Africa, Sweden, United States.

11 A selection of World Standard Population data are available from the WHO website (https://apps.who.int/healthinfo/statistics/mortality/whodpms/definitions/pop.htm).

12 Spatially disaggregated (county-level) population weights are reported in Colvin et al. (2021).

13 Walsh (1970) compares the registration of births with census reported births and notes that registration improves significantly by the 1910s.

14 The same method is used for sub-national population projections by the statistical agencies of the Republic of Ireland (https://www.cso.ie/en/methods/surveybackgroundnotes/populationandlabourforceprojections) and Northern Ireland (https://www.nisra.gov.uk/publications/2018-based-population-projections-areas-within-northernireland).

15 These are mostly return migrants from emigrant destination (see Fernihough and Ó Gráda 2019). Ireland traditionally experienced net emigration, the main reason for sustained population decline over time. Pre-war immigration was 20% of the emigration total. We assume immigration follows a similar spatial pattern as emigration.

16 Census figures provide details of location of birth of the population. We can estimate internal migration by comparing the 1901 and 1911 censuses (BPP 1901; 1913a). In 1901, 11.38% of the population of counties was born outwith the county; in 1911, this figure rose to 13.20%. This equates to a growth of 0.18% per annum. The outliers in this were the major urban centres (Belfast and Dublin), with the lowest rate of internal migration seen in the west (Kerry, with 3.85% in 1901 and 3.96% in 1911). Internal migration tended to be to the nearest urban centre, so province-rather than county-level statistics essentially already incorporate internal migration.

17 This census was conducted separately for the by then jurisdictionally partitioned island (Government of Northern Ireland 1929; Roinn Tionscail agus Tráchtála 1928, 1929).

18 See Blum et al. (2017) for a discussion of age heaping in Irish census data.

19 Our enlistment figure is considerably lower than Bowman’s (2014) recent estimates, who believes 210,000 Irish served in the British Army. This discrepancy is because the Army figures of enlistment for Ireland explicitly exclude ‘Irishmen enlisted in Great Britain who came over for the purpose’ (BPP 1921a, p. 6).

20 Bowman’s (2014) estimated mortality is 5.7% of all reported UK casualties, although his enlistment estimates suggest Irish comprised 4% of the British Army, implying Irish soldiers were over-represented in mortality figures.

21 The average difference between the sum of our county population estimates and those estimated by the Registrar General is −0.26%. We attribute this difference to our treatment of troop movements.

22 A similar argument can be made for Covid-19 mortality statistics, which we think should be consistently reported for the over-80s alongside the cruder measures typically used by journalists.

23 Indeed, it is these linear interpretations of population that are used in recent economic studies of the Spanish flu (e.g., Correira et al. 2020).

24 At a national level the linear interpolation using the 1901-1911 population growth is close 0.15% per annum, versus 0.18% for 1911-1918 using the component method.

25 Take two examples: Leitrim’s growth rate between 1901-1911 was −0.86% per annum, while the population growth was effectively zero between 1901-1918 owing to a reduction in emigration. Galway also experienced negative growth between 1901 and 1911 (−0.55%), but experienced growth between 1911 and 1918 (0.15%) owing to falling emigration.

26 Comparison of crude death rates shows regions today that have undergone the second demographic transition are 50-60% of those in the 1910s (Lesthaeghe 2014). But such comparisons are somewhat misleading given changes to countries’ demographic composition; in the US, for example, age-adjusted death rates are considerably lower than crude death rates, and there are age-adjusted disparities between ethnic groups (see, e.g., Kochanek et al. 2019).

27 Noteworthy here is that 64 per cent of the reported influenza deaths in 1900 were over the age of 55. However, the 1900 influenza pandemic was confined to one quarter, during the winter flu season, whereas Influenza-18 had excess mortality over multiple quarters.

28 The over-55 share of the population was 10% in the US and 12% in England and Wales (BPP 1917a).

29 In our maps we report Dublin City as part of Dublin County, and Belfast as part of Antrim. Deaths attributed to these cities may otherwise over-report the true impact of the pandemic because the main hospital infrastructure for these cities’ immediate hinterlands was located there.

30 Note that intergenerational family units have also been found to associated with higher fatality rates for the Covid-19 pandemic (Aparicio Fenoll and Grossbard 2020).

31 This is disproportionately driven by infant mortality (under the age of one).

32 Another property of the male population is that men deemed physically unfit for military service are among those left behind. It is feasible this selected population was more at risk of dying in the pandemic.

33 The heightened disease exposure of the working population is consistent with evidence from the US on modern seasonal influenza virus transmission: Markowitz (2019) find that a one percentage point increase in the employment rate increases the number of influenza-related outpatient health care visits by 19%.

34 Our contribution somewhat relates to Aburto et al. (2021), who find that off-the-shelf demographic methods also help to improve our understanding of life expectancy during a pandemic.

35 Using influenza-related cause of death statistics yields an estimated death toll between 20,000 and 31,000 individuals, similar in magnitude to those of Marsh (2010, chap. 2). This represents about 0.7% of Ireland’s population – considerably lower than the European total of 1.1% reported by Ansart et al. (2009).

36 The timing of the first reported Covid-19 cases was very similar both north and south of the border: the first case in the North was on 27 February and in the South on 29 February. Both cases were from citizens returning from Italy via Dublin airport. The blanket “lockdown” differed in both polities: the South instigated a lockdown on 12 March, while the North imposed theirs more gradually, with the full raft of measures arriving on 28 March.

37 For example, an op-ed in *The Irish Times* uses crude death rates to argue that the North could learn from the South (Tomlinson 2020), when in fact age adjustments suggest the opposite may be true. This is an argument we have subsequently made in our own op-ed, also published by *The Irish Times* (Colvin and McLaughlin 2020b).

38 The latest available data count 31,999 registered beds in the Republic of Ireland and 16,007 in Northern Ireland, while the population over 65 is 357,700 in the Republic and 168,764 in the North (HIQA 2020, RQIA 2018).

39 Especially the role of “blanket” Do Not Resuscitate orders (see discussion in QNI 2020).

